# Refining the diagnosis of Gestational Diabetes Mellitus: A systematic review to inform efforts in precision medicine

**DOI:** 10.1101/2023.04.19.23288627

**Authors:** Ellen C. Francis, Camille E. Powe, William L. Lowe, Sara L. White, Denise M. Scholtens, Cuilin Zhang, Yeyi Zhu, Marie-France Hivert, Soo Heon Kwak, Arianne Sweeting

## Abstract

**Background:** Among people with gestational diabetes mellitus (GDM), there is inter-individual variability in clinical outcomes that appears to be related to factors beyond glycemia. However, the precise factors (information on the unique pathophysiology within a person, environment, and/or context) that may help refine the diagnosis of GDM remain unclear. To determine if a precision medicine approach could refine the diagnosis of GDM, we conducted a systematic review of a variety of potential precision markers analyzed in studies among individuals with GDM.

**Methods:** Systematic literature searches were performed in PubMed (https://pubmed.ncbi.nlm.nih.gov/) and EMBASE (https://www.embase.com) databases from inception to March 2022 for observational studies and controlled trials. Studies were included if they reported data on, and compared outcomes between, individuals with GDM. The following categories of precision markers were included in the current search: non-glycemic biochemical markers (cholesterol, insulin profiles); genetics/genomics or other -omics (proteomics, lipidomics, metabolomics, metagenomics); maternal/fetal anthropometric (eg., maternal BMI, gestational weight gain, fetal biometry ultra-sound measures); clinical risk factors (medical/familial history, prior delivery complicated by macrosomia or a large for gestational age [LGA] neonate); sociocultural or environmental modifiers (diet, smoking, race/ethnicity, socioeconomic status).

**Results:** We focused on synthesizing the literature on genetics, -omics, non-glycemic biomarkers, maternal anthropometry/fetal biometry, and clinical/sociocultural risk factors. A total of 5,905 titles and abstracts were screened, 775 underwent full-text review, and 137 studies that included a total of 432,156 GDM cases were synthesized. Of the studies on non-glycemic biomarkers (n=33), lipids and insulin sensitivity/secretion indices were the two most common precision markers, with elevated maternal triglycerides and insulin resistance generally associated with greater risk of LGA and macrosomia. Studies of genetics or other -omics were scarce (n=5); however, differences in genetic variants in adiponectin or adiponutrient genes and non-coding RNAs accounted for variability in perinatal outcomes. The majority of studies (n=77) evaluated maternal anthropometry or fetal biometry as a precision marker, and these studies demonstrate that individuals with adiposity who develop GDM are at a substantially higher risk of LGA or macrosomia than those with GDM and lower adiposity. There were 49 studies evaluating GDM risk factors or sociocultural markers, with only six studies examining multiple risk factors as a composite marker. There were inconsistent findings that GDM risk factors, such as older maternal age, accounted for variation in adverse outcomes.

**Conclusions:** Our review demonstrates that a major gap exists in studies examining non-glycemic biochemical, genetic, or other ‘omic precision markers among individuals with GDM. Given that people meeting current diagnostic criteria for GDM may have different risk profiles, our review identifies several factors (including obesity, insulin resistance, hypertriglyceridemia) that may be useful in risk stratification of GDM, setting the stage for a precision approach to its diagnosis.

## INTRODUCTION

Gestational diabetes (GDM) is the most common metabolic complication of pregnancy with an increasing prevalence consistent with the concomitant global increase in obesity and diabetes^1^. GDM is traditionally referred to abnormal glucose tolerance with onset or first recognition during pregnancy, which is typically tested for between 24-28 weeks’ gestation^2^. It is associated with increased maternal and neonatal complications such as hypertensive disorders of pregnancy and preeclampsia, birth trauma, neonatal respiratory distress, neonatal hypoglycemia, and macrosomia^3^.

It is important to note that not all cases of GDM carry the same risk of adverse outcomes. While the diagnostic criteria for GDM focus on detecting dysregulation of glucose metabolism, GDM is a disorder of all metabolic fuels, and is increasingly recognized as a heterogeneous condition^4, 5^. Several upstream determinants of metabolic health are considered risk factors for the development of GDM, including higher body mass index (BMI) as well as sociocultural factors. Both clinical and metabolic differences among individuals with GDM may modify the impact of the condition on maternal and fetal health^6^. However, efforts to systematically review studies evaluating clinical and sociocultural/environmental risk factors, genetics, -omics and non-glycemic biomarkers that could identify subgroups within GDM with higher risk of adverse perinatal outcomes are lacking. In an era with increased attention to precision medicine, this lack of systematic data represents a critical gap. Therefore, it is essential to characterize markers that modify the effect of GDM on adverse perinatal outcomes to inform whether the diagnosis can be refined. This knowledge may lead to different clinical actions during pregnancy for different GDM subtypes and argue for the development of novel interventions to reduce adverse pregnancy and perinatal outcomes.

The Precision Medicine in Diabetes Initiative (PMDI) was established in 2018 by the American Diabetes Association (ADA) in partnership with the European Association for the Study of Diabetes (EASD). The ADA/EASD PMDI includes global thought leaders in precision diabetes medicine who are working to address the burgeoning need for better diabetes prevention and care through precision medicine^7^. As part of the ADA/EASD PMDI effort, we aimed to review the existing literature to investigate GDM subtypes and heterogeneity among GDM in association with adverse perinatal outcomes. This effort was undertaken to aid in determining whether a precision medicine approach could refine the diagnosis of GDM beyond traditional glycemic measures.

## METHODS

A protocol for this review was registered at PROSPERO (CRD42022316260) on 11 March 2022.

### Data sources and search strategy

Systematic literature searches were performed in PubMed (https://pubmed.ncbi.nlm.nih.gov/) and EMBASE (https://www.embase.com) databases from inception to March 2022 for observational studies and controlled trials that reported data on, and compared outcomes between, individuals with GDM. The following categories of precision markers were included in the current search: non-glycemic biochemical markers (cholesterol, insulin profiles); genetics/genomics or other -omics (proteomics, lipidomics, metabolomics, metagenomics); maternal/fetal anthropometric (eg., maternal BMI, gestational weight gain, fetal biometry ultra-sound measures); clinical risk factors (medical/familial history, prior delivery complicated by macrosomia or a large for gestational age [LGA] neonate); sociocultural or environmental modifiers (diet, smoking, race/ethnicity, socioeconomic status). The search was restricted to studies in adult humans, published in English. The search strategy is available in **Supplementary Material 1.**

### Selection criteria

We included studies that included at least 100 participants and 30 GDM cases. Study outcomes needed to be reported among GDM cases or compared between subgroups of participants with GDM. Studies that reported on one or more common pregnancy and perinatal outcomes related to GDM diagnosis were included. Maternal outcomes included hypertensive disorders in pregnancy, preeclampsia and cesarean delivery. Offspring outcomes included anthropometry at birth (macrosomia, LGA offspring, small-for-gestational-age [SGA] neonate), preterm delivery, birth trauma, metabolic sequelae or mortality. Studies evaluating prevention, prediction, treatment, long-term maternal and offspring outcomes or glycemic markers to risk stratify or subgroup individuals with GDM were the objective of complementary reviews, or were beyond the scope of the present review.

As our main goal was to review studies that were offering GDM subtyping beyond glycemia, we excluded studies that only reported on glycemic based biochemical markers (e.g., HbA1c, fasting glucose, oral glucose tolerance test glycemic thresholds). We also excluded studies that measured the precision marker after GDM diagnosis, did not report outcomes specifically among participants with GDM or GDM subgroups, studies which included overt diabetes (based on non-pregnancy glycemic thresholds) with GDM cases, studies among multi-gestations, or studies missing full-text or without full-text in English. We excluded studies that used total gestational weight gain (GWG) over the whole period pregnancy, or fetal biometry only after 32 weeks of gestation because these factors would not be suitable as a precision marker at or around the time of GDM diagnosis. All studies were screened by reviewers in duplicate. All titles and abstracts were screened for eligibility, and those that were assessed as potentially meeting inclusion/exclusion criteria were selected for full-text evaluation.

### Data extraction and quality assessment

Study and sample characteristics were extracted in duplicate from full-text using a web-based collaboration software platform that streamlines the production of systematic literature reviews (Covidence systematic review software, Veritas Health Innovation, Melbourne, Australia). The following data elements were extracted from each study when available: cohort characteristics (continent, country, study type [hospital/registry/cohort], enrollment years); participant characteristics (age, BMI, the proportion nulliparous); GDM information (GDM sample size, GDM diagnostic criteria or description); timing of precision marker measurement (pre-pregnancy, before or at GDM diagnosis); perinatal outcomes (maternal, fetal/neonatal). The risk of bias and overall quality of each study was assessed independently or in duplicate using the Joanna Briggs Institute Critical Appraisal Tool for cohort studies, which was modified specifically for the objectives of the current systematic review^8^ (**Supplementary Material 2)**. We assessed the studies using a ten-question measure and considered studies with two poor quality metrics to be of low quality.

### Data synthesis and analysis

For each category of precision marker, two independent reviewers jointly summarized the findings. Most studies reported outcomes among individuals with GDM for more than one precision marker (e.g. maternal BMI, also reporting on maternal age, prior history of GDM, etc.).

## RESULTS

### Literature search

The literature search yielded 5905 non-duplicated abstracts (Figure 1). After independent review by 2 investigators for each abstract, 5130 abstracts were excluded. Among the 775 full-text studies reviewed, 638 were excluded based on our study selection criteria (precision variable measured after diagnosis, no outcome reported among GDM cases, or because they were unrelated to the scope of the present review (**Figure 1**). After final exclusions, 137 studies met the inclusion criteria and were summarized in the present systematic review. The studies were categorized into three groups of precision markers 1) biochemical, genetics, ‘omics precision markers; 2) maternal anthropometry/fetal biometry; and 3) clinical risk factors, sociocultural or environmental modifiers.

**Figure 1:**
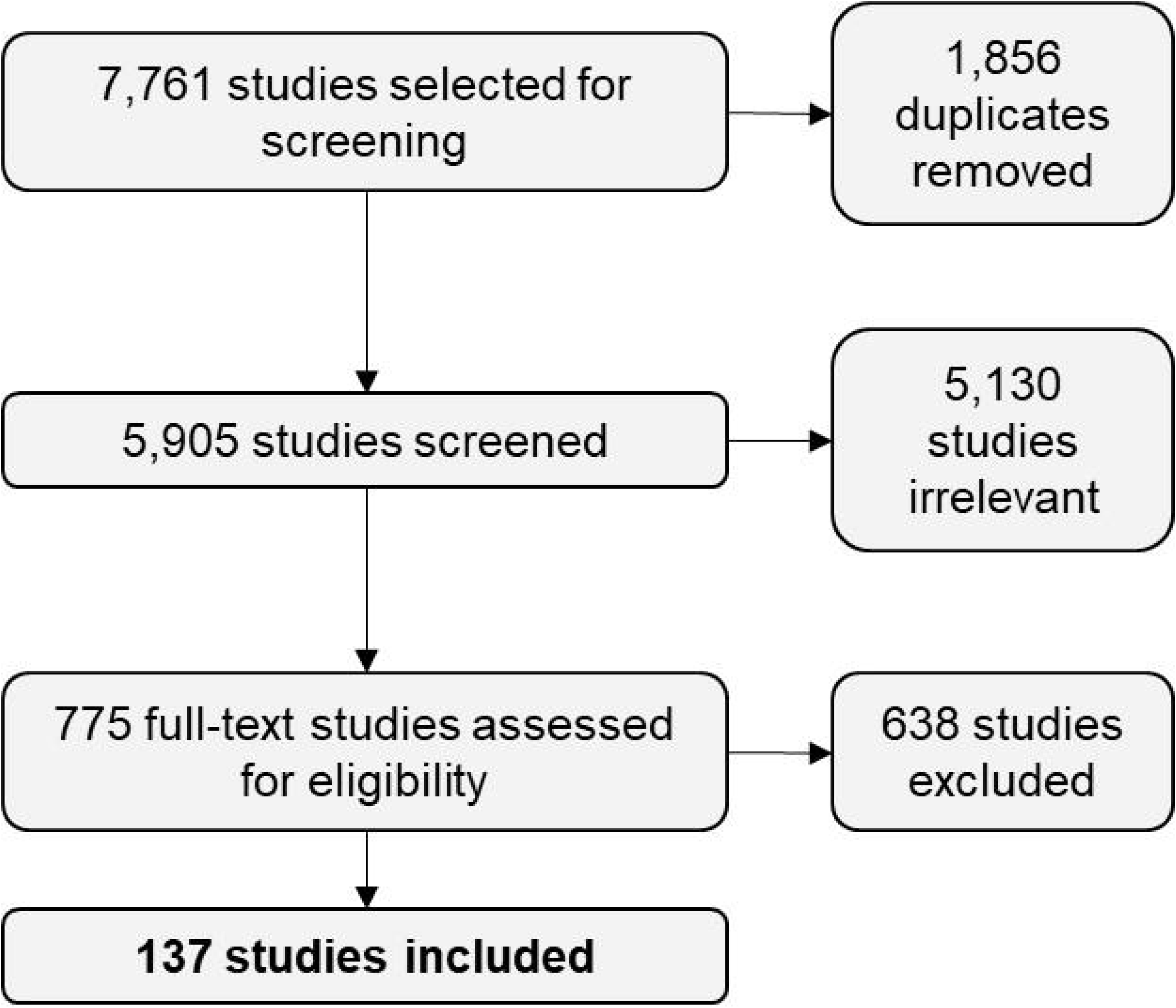
Article selection, screening, exclusion, and inclusion

### Overall study characteristics

Detailed study characteristics of the 137 studies representing a total of 432,156 participants are shown in **Table 1**. The median (range) number of participants was 587 (60 - 170,572). Of these studies, 33 evaluated non-glycemic biomarkers among individuals with GDM and five studies included genetic or ‘omic markers. The majority of studies (n=73) evaluated maternal anthropometry as a precision marker. There were 49 studies evaluating maternal clinical risk factors or sociocultural markers, with six studies examining multiple risk factors as a composite marker or algorithms/nomograms. Most studies (72%) included pregnancies from 2000-2020 and were from geographically diverse regions. Of the studies included, 20% were conducted in China, 12% in the USA, 7% in Australia, and 6% in Spain. The most frequent diagnostic criteria for GDM were either current IADPSG or WHO criteria.

**Table 1:**
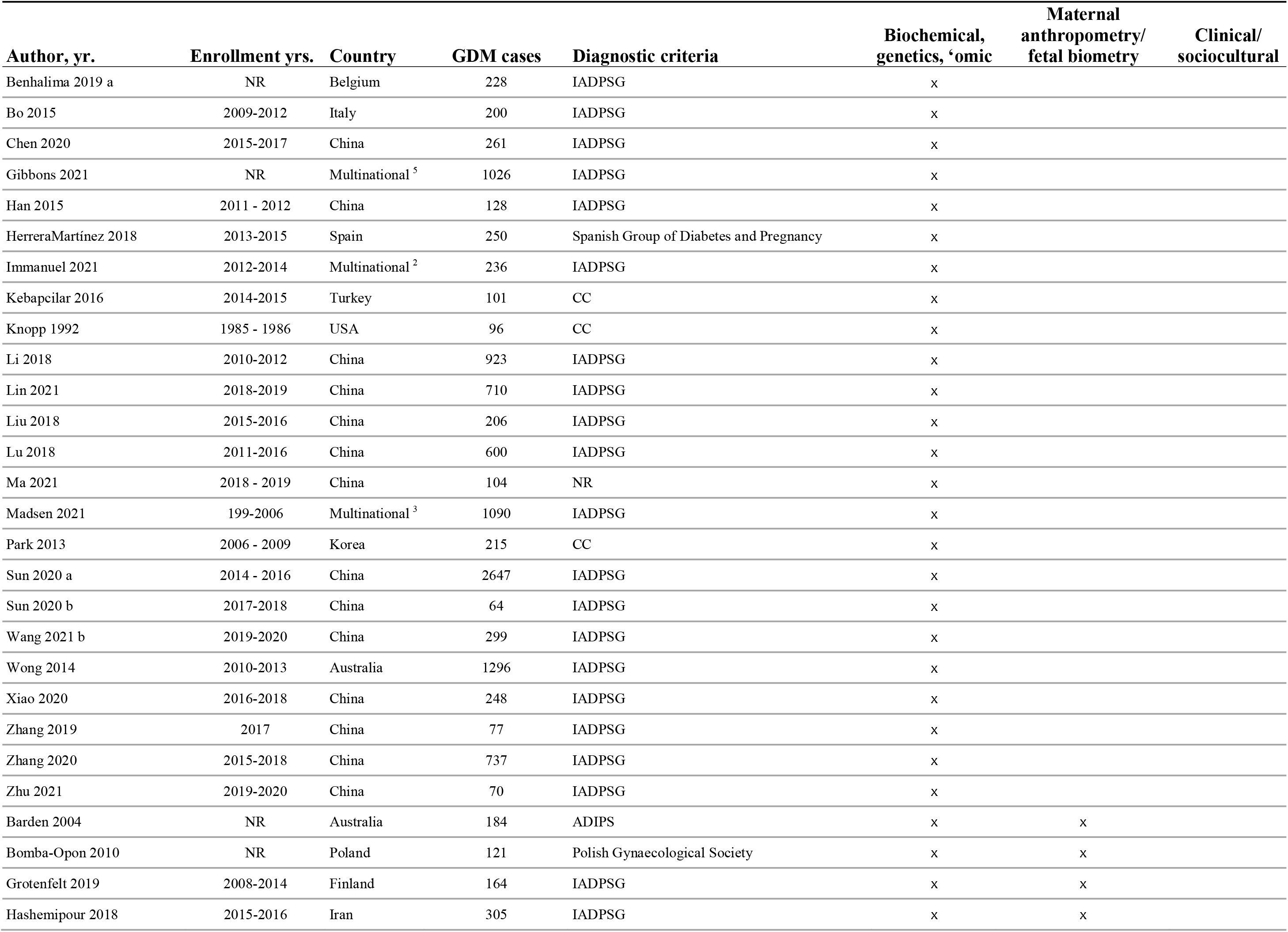
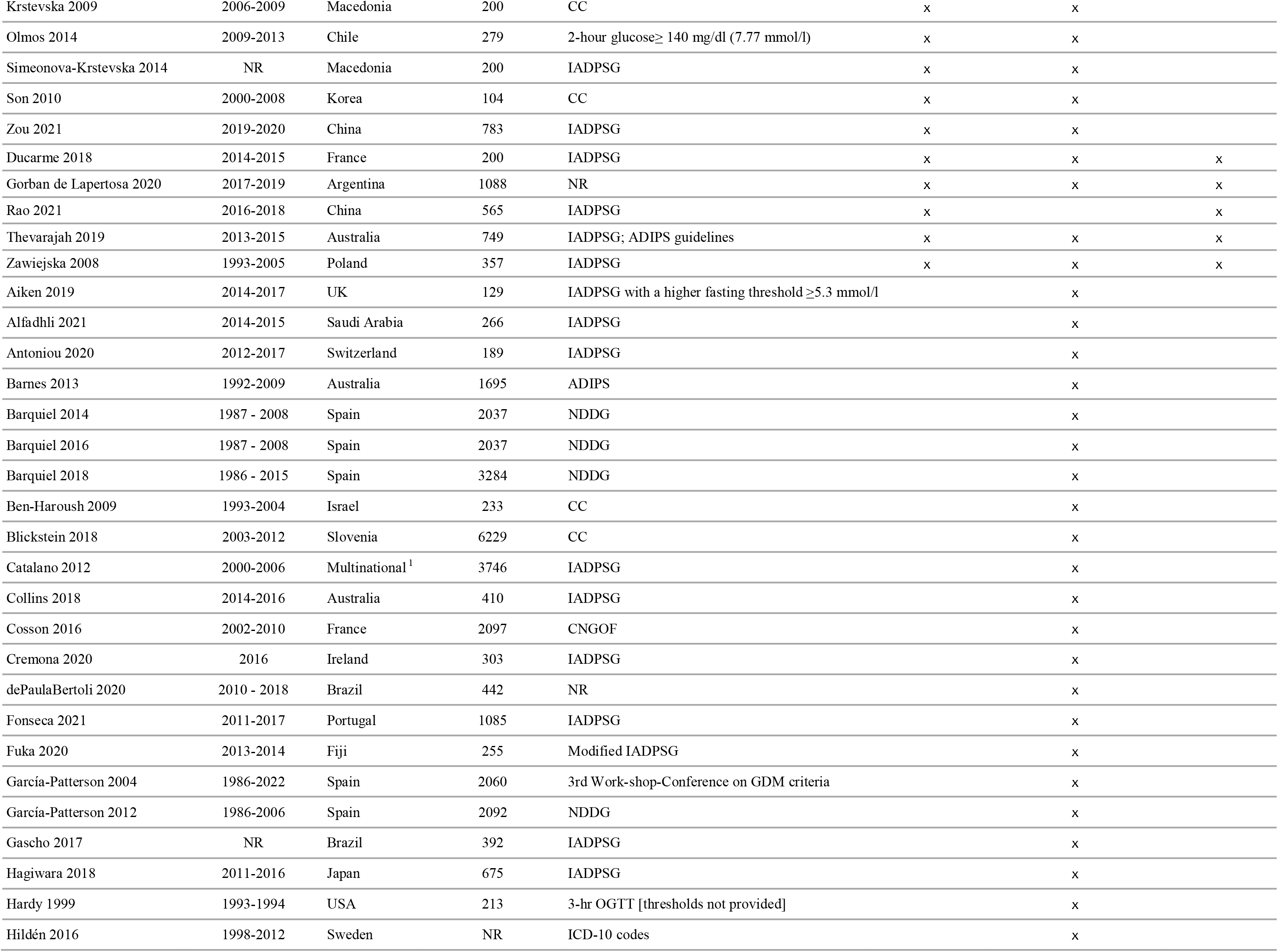
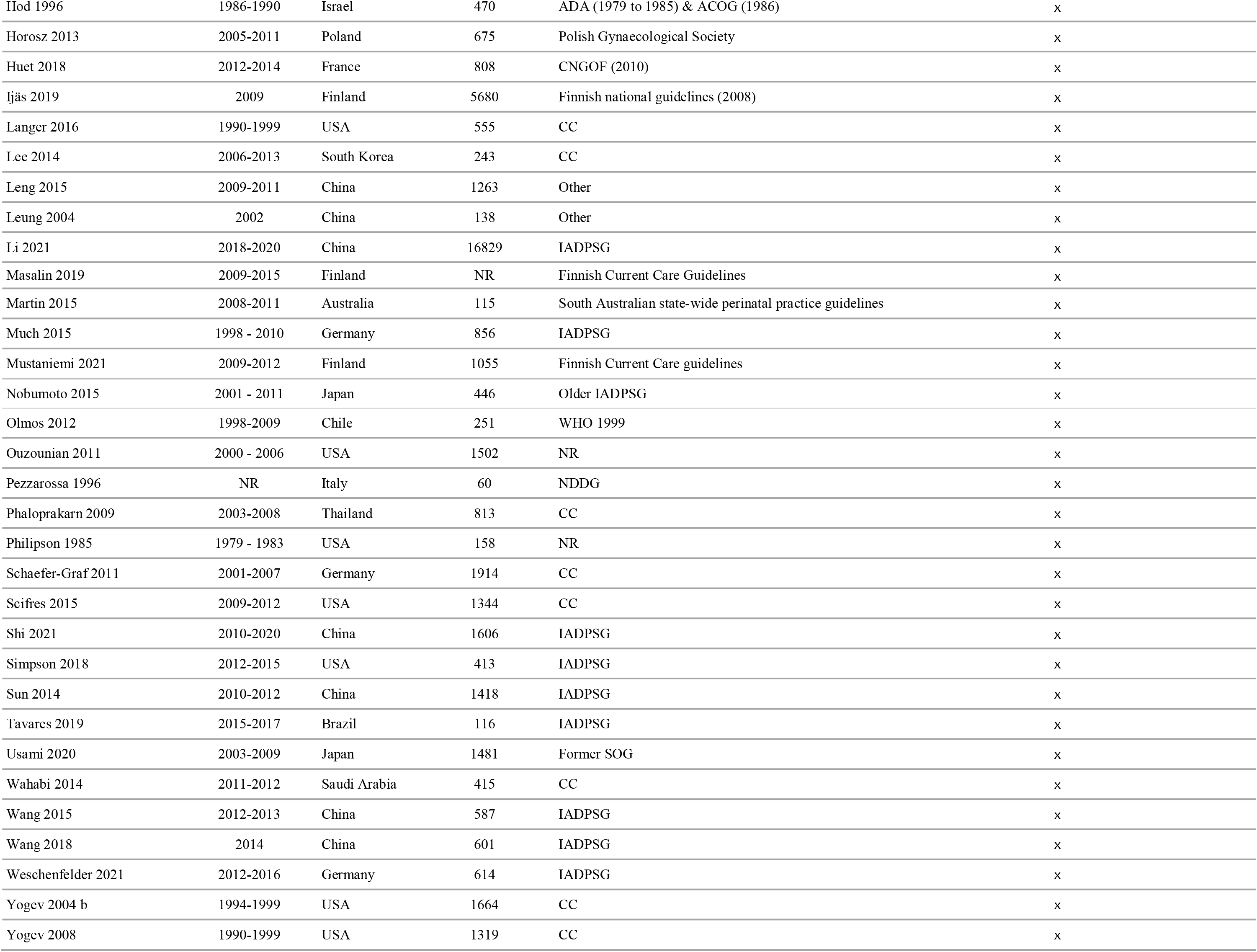
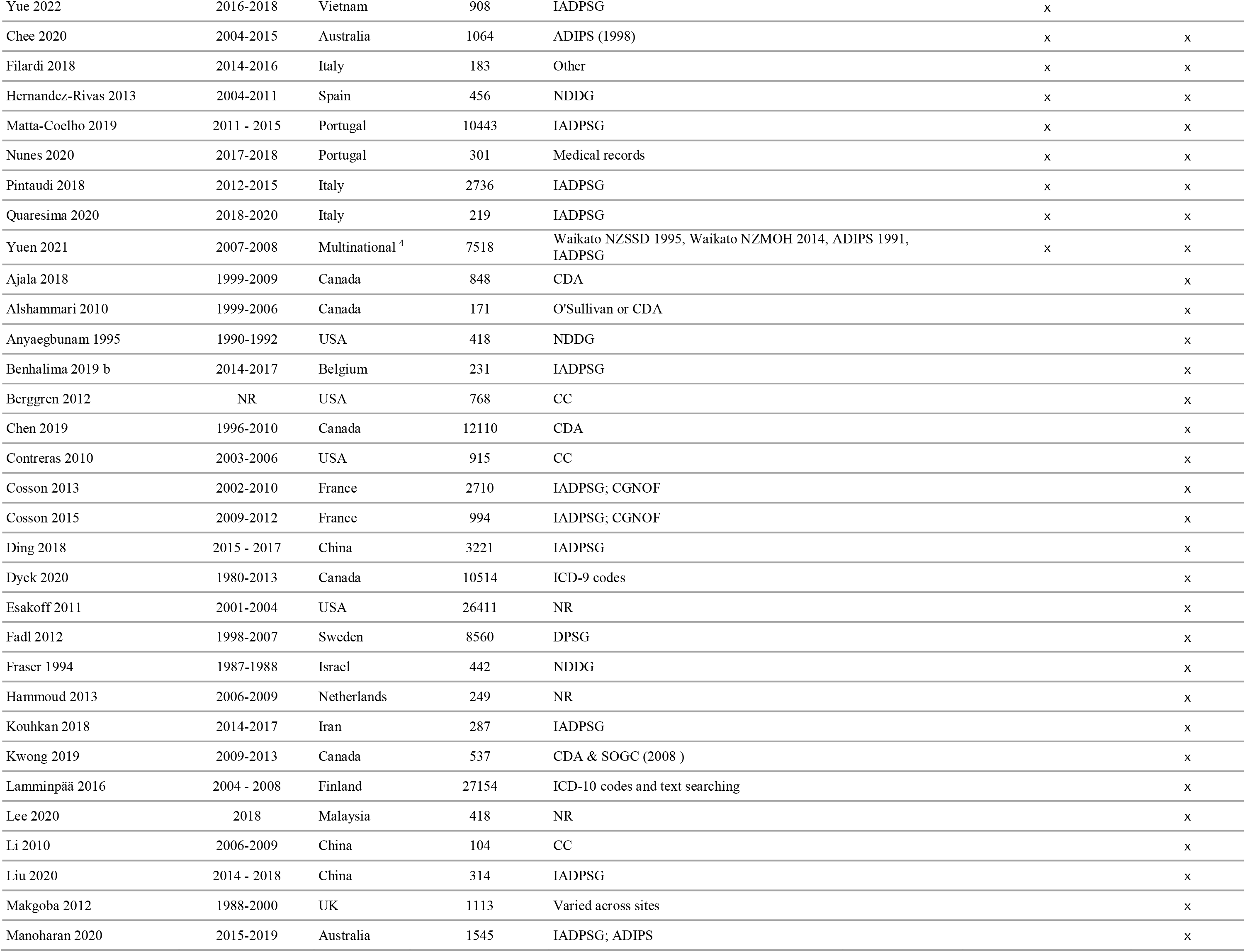
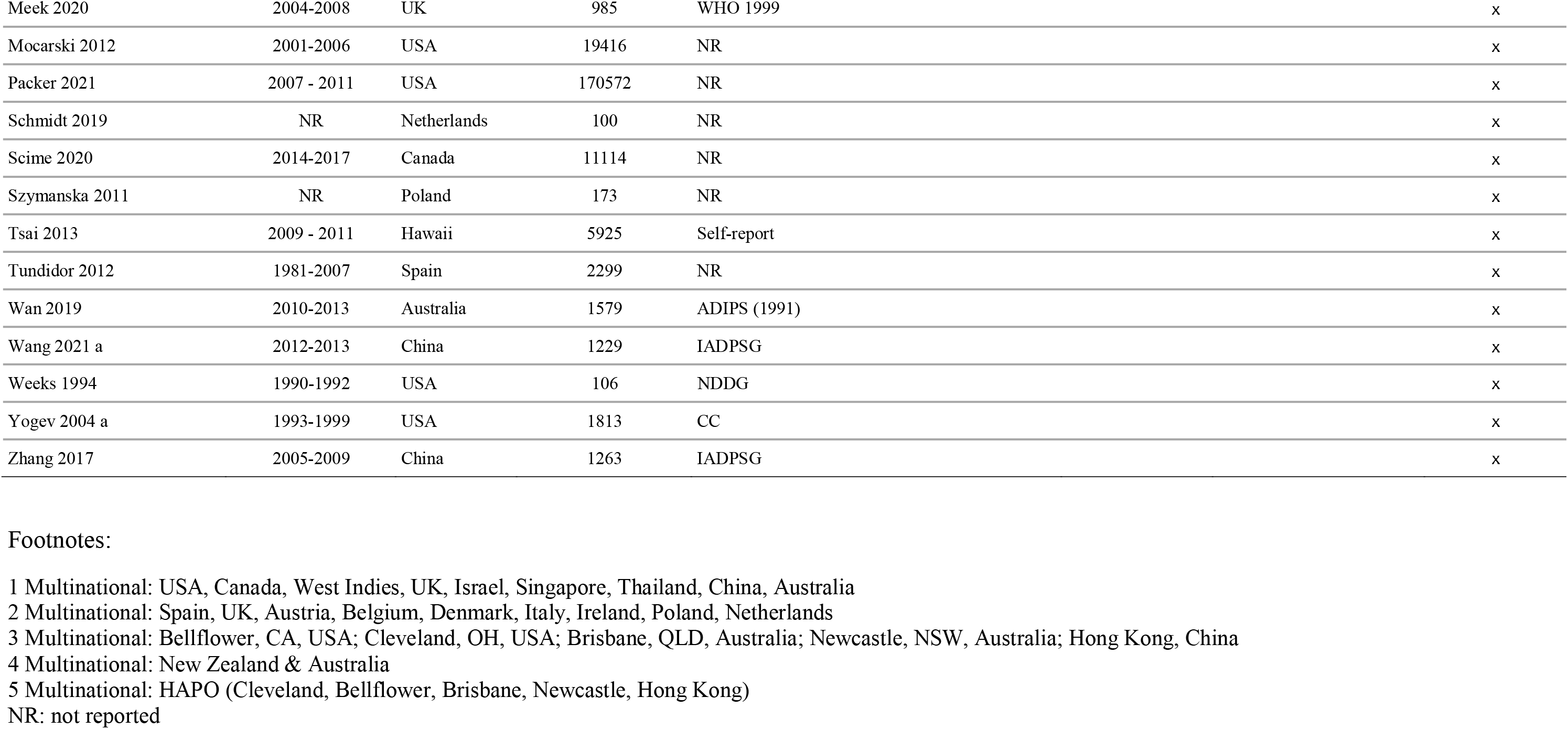
Study characteristics

Overall, 45% of the studies were considered to be of low quality (**Figure 2**).

**Figure 2:**
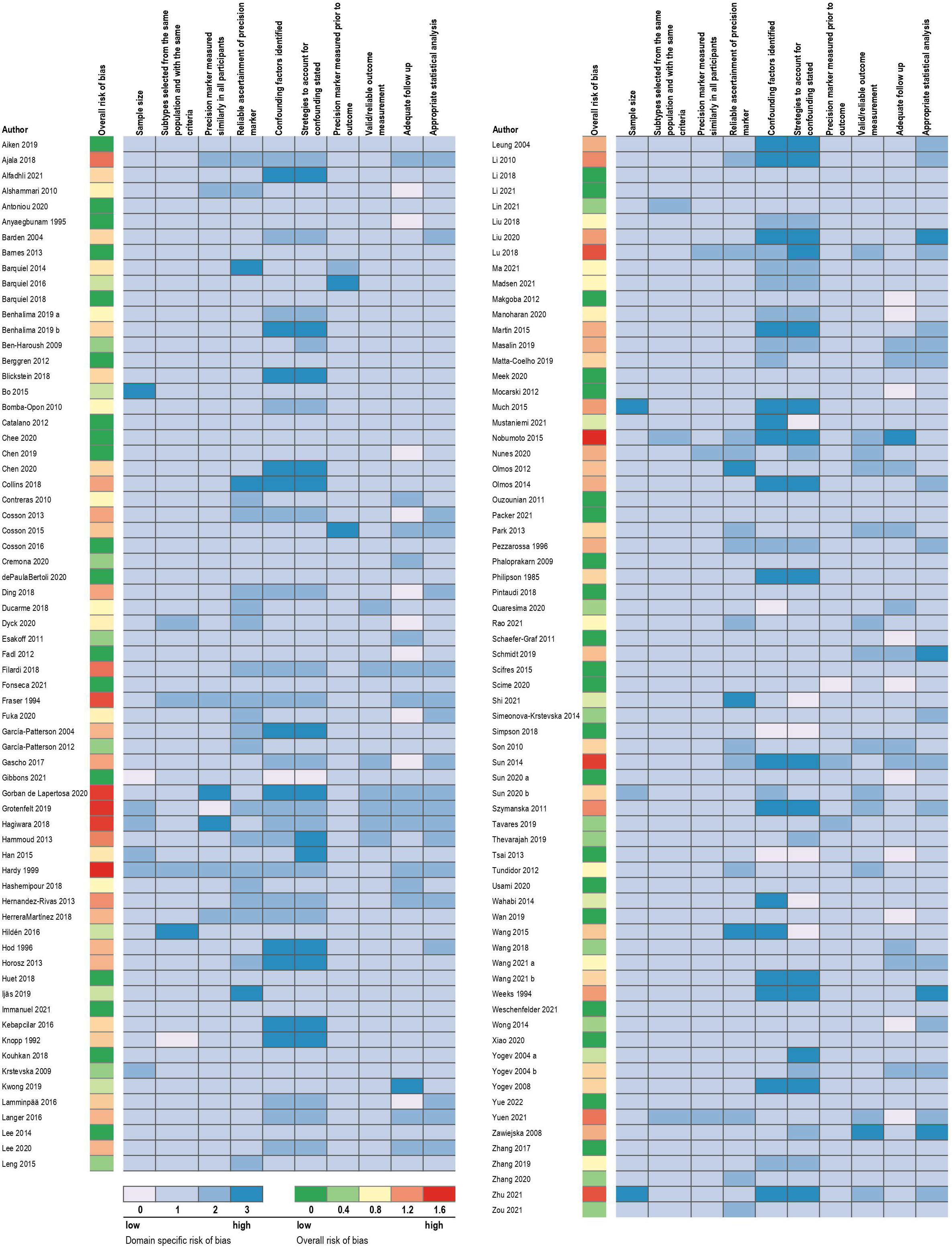
Quality assessment of each individual study overall and by critical appraisal domain

Approximately 40% of studies reported unadjusted estimates and therefore were ranked low on domains of confounding. Self-reported data is generally considered to be of low quality, and since most studies included self-reported pre-prepregnancy weight, 28% of studies were ranked as poor on the ascertainment of precision marker domain. Other factors that impacted the quality rankings were mostly due to unclear reporting in the manuscripts.

### Biochemical, genetics, ‘omics precision markers

#### Study characteristics

A total of 38 unique studies reported associations of biochemical, genetics, or other -omics markers with adverse pregnancy and perinatal outcomes among participants with GDM. Of these studies, 15 described associations of lipid classes (triglycerides, total cholesterol, LDL cholesterol, and HDL cholesterol) with adverse pregnancy and perinatal outcomes^9–23^. There were 12 studies that described associations of insulin sensitivity/resistance profiles^13, 24–30^, or insulin secretion or insulin dynamic indices^9, 31–33^ with perinatal outcomes. A small number of the included studies subtyped GDM based on adipokines (n=2)^31, 34^, metabolomics (n=1)^35^, non-coding RNA (n=2)^36, 37^, and candidate gene studies (n=2)^38, 39^. The detailed characteristics of these studies are summarized in **Supplementary Table 1,** which also includes a few studies with measurement of less precise biochemical markers (eg, proteinuria, platelet count). Most studies (63%) included pregnancies from 2010-2020, diagnosed GDM using IADPSG criteria (55%), and included a median (range) number of GDM cases of 242 (64-2,647). Most studies were of medium to high quality, which was attributable to the reporting of unadjusted estimates and therefore being ranked lower when assessing whether the study accounted for confounding. (**Supplementary Figure 1**).

#### Lipid subclasses

Among the 14 studies measuring triglycerides prior to or at the time of GDM diagnosis^9–11, 13–23^, approximately half reported that higher triglycerides were positively correlated with increased birthweight or risk of LGA or macrosomia^17–23^, whereas the other studies were null^9–11, 13–16^. Of the studies measuring total cholesterol, LDL, or HDL cholesterols (n=12)^10–16, 18, 20–23^, one found a positive association of LDL with LGA^21^, and three reported lower mean HDL levels among individuals who later had an LGA baby^12, 14, 15^.

#### Insulin profiles and indices

A variety of methods of calculating insulin resistance/sensitivity and insulin secretory dysfunction using timed insulin and glucose concentrations measured through the OGTT for GDM subgroup stratification were described. The homeostatic model assessment of insulin resistance (HOMA-IR or HOMA2-IR) calculated using fasting insulin and glucose levels at the time of GDM diagnosis (http://www.dtu.ox.ac.uk/homacalculator/)^40^ was used most commonly. The Matsuda index^41^, modelled using fasting glucose and insulin values across the OGTT, and HOMA-S (modelled using fasting glucose and insulin)^42^, were the most frequent measures of insulin sensitivity. HOMA-B/HOMA-2B (http://www. dtu.ox.ac.uk; homeostatic model assessment of beta-cell function; fasting insulin and fasting glucose model) and the Stumvoll first phase insulin estimate (modelled using timed insulin and glucose values from OGTT)^43^ were the most utilized indices defining insulin secretory function. Other indices such as the insulinogenic index and disposition index were utilized rarely^24, 27^.

Among studies calculating HOMA-IR, all four found that individuals with GDM and high HOMA-IR (highest quartile or >2.0) had a significantly increased risk of LGA or macrosomia^13, 25, 29, 33^, although in one study the statistical comparison was to normal glucose tolerant individuals. In two studies among GDM only, insulin profiles such as a defect in insulin sensitivity, insulin secretion, or a combination of both were not associated with differences in perinatal outcomes^26, 28^. Three studies reported on insulin profiles among participants with and without GDM^24, 27, 30^. In two of these, participants with GDM and defects in insulin sensitivity or a combination of both defects had higher rates of LGA and macrosomia (where GDM individuals with isolated insulin secretion defect were at similar risk to individuals without GDM); however, there were no reported tests to assess whether these rates were statistically different only among the GDM cases^27, 30^. A study of insulin secretion peaks during an OGTT found that a delayed insulin secretion peak was associated with increased risk of preeclampsia, LGA, and neonatal hypoglycemia^32^, whereas both a study of insulin following a 50g glucose load and a study of fasting plasma insulin, found no association with adverse perinatal outcomes^9, 31^.

#### Adipokines

Two studies measured adiponectin, leptin^31, 34^, and one additionally measured visfatin^34^. Neither study found that adiponectin or leptin were associated with perinatal outcomes; however, higher visfatin levels were associated with lower risk of LGA^34^.

#### Metabolomics

A single study utilizing mass spectrometry examined the association of plasma levels of carnitine and 30 acylcarnitines with newborn complications in individuals with GDM^44^. Carnitine and three acylcarnitines were associated with GDM after adjustment for covariates. Carnitine and acylcarnitine levels together with clinical factors were used to construct a nomogram to predict macrosomia within the GDM group, which resulted in an area under the ROC curve of 0.78

#### non-coding RNAs

Two studies examined the association of different classes of non-coding RNAs with various adverse pregnancy outcomes^36, 37^. One study of circulating long non-coding RNAs (lncRNAs) measured in 63 GDM cases found that including XLOC_014172 and RP11-230G5.2 in a prediction model for macrosomia resulted in an area under the receiver operator characteristic curve of 0.962^36^. In a study of high or low plasma levels of circular RNA circATR2, high circATR2 was associated with higher rates of prematurity, miscarriage, intrauterine death, fetal malformations, intrauterine infection and hypertension but not macrosomia or fetal distress^37^.

#### Candidate gene studies

Two studies used a candidate gene approach to subtype individuals with GDM based on their genotype and examine associations with pregnancy outcomes^38, 39^. One study of a variant in the patatin-like phospholipase-3 (PNPLA3)/adiponutrin gene (rs738409 C.G), found that G allele (n=96) vs. CC homozygotes (n=104) was associated with higher levels of AST, ALT, and GGT and lower fasting insulin, insulin resistance and LGA birth^39^. In a study of SNP 45TG in exon 2 of the adiponectin gene, the G allele and GG + TG genotypes were associated with GDM, lower adiponectin levels and greater incidence of macrosomia and neonatal hypoglycemia compared to the TT group^38^.

### Maternal anthropometry or fetal biometry precision markers

#### Study characteristics

A total of 77 unique studies reported associations of maternal anthropometric or fetal biometry ultra-sound measures. Of these, 68 described associations of pre-pregnancy overweight and obesity defined by maternal BMI (>25.0-29.9 kg/m^2^ and ≥30.0 kg/m^2^, respectively) with adverse perinatal outcomes. A small number of studies described the relationship of early gestational weight gain (early GWG) prior to diagnosis (n=4)^45–48^, or fetal biometry ultra-sound measures (biparietal, head, abdominal circumference or femur length) (n=9)^49–56^ with adverse perinatal outcomes. The detailed characteristics of these studies are summarized in **Supplementary Table 2**. Studies most frequently (40%) included pregnancies from 2010-2020 and diagnosed GDM using IADPSG, or modified IADPSG criteria (41%). The median (range) number of GDM cases was 594 (60-16,829). Most studies were of medium to high quality, which was attributable to the reporting of unadjusted estimates and therefore being ranked lower when assessing whether the study accounted for confounding (**Supplementary Figure 2**).

#### Body Mass Index

Studies evaluating the relationship between maternal BMI and adverse pregnancy outcomes tended to be retrospective hospital record cohort or case control studies relying on self-reported pre-pregnancy weight. Of the 68 studies, all but nine^10, 13, 20, 23, 30, 49, 50, 53, 56–59^ reported positive associations between maternal overweight and obesity and adverse perinatal outcomes.

The most consistently reported association was observed for maternal pre-pregnancy obesity and neonatal birthweight, LGA and macrosomia. Regarding maternal pre-pregnancy overweight and obesity as a precision factor, numerous studies showed associations with greater risk of cesarean delivery^56, 59–69^ or preeclampsia/hypertensive disorders of pregnancy^16, 61, 63, 64, 67, 70–75^. Four studies reported an association with neonatal hypoglycemia^76–79^, two studies reported an association with a composite outcome of neonatal morbidity and/or admission to NICU^58, 70^, and one study reported an increased risk of major congenital malformations^80^.

#### Early gestational weight gain

Three of four^45–48^ studies of early GWG in individuals with GDM found positive associations with LGA^45, 47, 48^, one of which reported that trimester-specific weight gain above the Institute of Medicine Guidelines was additionally associated with increased risk of preeclampsia and macrosomia^45^.

#### Fetal biometry

Among the studies with a fetal biometry ultra-sound measure near the time of GDM diagnosis, six of the eight studies found that larger fetal abdominal^81–85^ or biparietal circumference^22^ was positively associated with greater neonatal size (birthweight, LGA, macrosomia).

### Clinical risk factors, sociocultural and environmental measures

#### Study characteristics

A total of 49 unique studies reported associations of individual clinical or sociocultural risk factors, or associations of multiple risk factors with adverse perinatal outcomes among individuals with GDM. Of these studies, six included multiple risk factors as a composite variable^66, 86–90^. Eight studies among individuals with GDM examined unique risk factors (e.g., fetal sex, seasonality of conception, assisted reproduction)^91–97^. The majority (n=35) of studies tested associations of factors that increase risk of GDM in the general population, such as family history of diabetes, older age, and higher maternal BMI, with adverse perinatal outcomes among individuals with GDM. The detailed characteristics of these studies are summarized in **Supplementary Table 3.** Half of the studies included pregnancies from 1990-2009, and four studies from the 1980s. A third of studies diagnosed GDM using IADPSG, or IADPSG modified criteria, and 20% did not report diagnostic criteria. The median (range) number of GDM cases in these 50 studies was 950 (100-170,572). Most studies were of medium to high quality, which was attributed to the reporting of unadjusted estimates and therefore being ranked lower when assessing whether the study accounted for confounding (**Supplementary Figure 3**).

#### Composite of multiple clinical or sociocultural risk factors

Studies examining multiple clinical or sociocultural risk factors often included data from medical/familial history (prior GDM pregnancy, family history of diabetes), maternal/fetal anthropometry (pre-pregnancy obesity, prior LGA/macrosomia delivery). Four studies found that GDM with one or more risk factors was associated with greater neonatal size (birthweight percentile, LGA, macrosomia), compared to GDM without risk factors^66, 86–88^, with two of these studies also finding a higher risk of cesarean delivery^66, 88^. One study reported that GDM with one or more risk factors was associated with cesarean delivery and not neonatal size^90^, and another found no difference in perinatal outcomes among individuals with or without risk factors^89^.

#### Individual clinical or sociocultural risk factors

Individual risk factors such as maternal age, race, polycystic ovarian syndrome, parity, prior history of GDM, prior history of macrosomia, and/or family history of diabetes were often reported in studies with a primary analysis focused on other precision factors (e.g. maternal BMI, biomarkers). However, these risk factors were not the focus of our review, thus we only summarized general observations from our literature assessment in **Supplementary Table 3**. Of note, in addition to studies reporting on GDM risk factors as a potential precision marker, there were four studies reporting on psychological factors (depression, anxiety, diabetes distress), and nine studies that examined unique clinical or sociocultural modifiers, which included markers such as fetal sex, seasonality of conception, assisted reproduction, all of which have been summarized in **Supplementary Table 3.** Two studies found that individuals with GDM and a history of PCOS were at higher risk of preeclampsia, and subsequent delivery of offspring with higher risk of SGA birth weight^98, 99^.

Studies have reported various findings from comparing outcomes in individuals with GDM from different races or ethnicities. Race is a social construct and is correlated with a variety of factors that are specific to social context including experiences of racism, some aspects of culture, socioeconomic status, and many other factors that may influence health outcomes. In the social context of United States (US), individuals with GDM who self-identified as African-American were at higher risk of perinatal complications, including fetal death^100, 101^, data mirroring health disparities leading to different perinatal complications rates in the general US population. Findings were inconsistent regarding risk of complications in individuals with GDM who identified as Hispanic (versus non-Hispanic): most studies did not find major differences in adverse outcomes^102, 103^ while one large study reporting higher rate of preterm birth^100^. In Hawaii, white individuals with GDM were more likely to give birth to baby with macrosomia compared to other race/ethnicity groups (Hawaiian/Pacific Islander, Filipina, or other Asian individuals)^104^. Several studies in Australia, USA, and Canada comparing individuals with GDM from different race/ethnicity found that individuals who identified as Asian were less likely to have babies classified as LGA (compared to a White-identified individuals)^101, 105–109^. In two Canadian studies, GDM participants from First Nations or Indigenous groups were at higher risk of perinatal complications^110^.

## DISCUSSION

Our systematic review of 137 studies and 432,825 individuals with GDM demonstrates that there is inter-individual variability in clinical outcomes that appears to be related to factors that extend beyond glycemia. Among individuals with GDM, those with higher triglycerides or markers of an insulin sensitivity defect (or high insulin resistance), are at higher risk of having a newborn classified as LGA or with macrosomia (overall moderate evidence level). Data from adjusted analyses suggest that this higher risk is only partially attributable to differences in pre-pregnancy BMI. Prior research has largely focused on the impact of pre-pregnancy adiposity on adverse perinatal outcomes. Based on studies of moderate quality, we found that the co-occurrence of adiposity and GDM was associated with an increased risk of LGA and macrosomia as well as related pregnancy complications (eg, cesarean delivery), compared to GDM without adiposity. Unsurprisingly, higher centiles of fetal biometry measured with ultra-sound in the 2^nd^ or 3^rd^ trimester were also precision markers for fetal overgrowth. There were inconsistent findings that GDM risk factors, such as older maternal age, accounted for variation in adverse outcomes.

Thus, further research is needed to assess whether the GDM risk factors commonly measured in the general population, are adequate to stratify risk of adverse perinatal outcomes within the subset of individuals who develop GDM. Below, we summarize our general findings and future directions for precision medicine in GDM diagnosis.

### Biochemical, genetics, omic precision markers

Most studies examining lipids among individuals with GDM in association with adverse perinatal outcomes have measured a standard lipid panel that includes three measures of cholesterol levels (total, LDL and HDL cholesterol) and triglycerides. Approximately half of the studies reported higher triglycerides were associated with macrosomia or LGA, with fewer studies finding that higher LDL or lower HDL was associated with neonatal size. Interestingly, the average pre-pregnancy BMI and the percentage of individuals with obesity appeared to be lower among the studies that found a positive association of triglycerides with neonatal size^17–23^ compared to the studies that reported no association null^9–11, 13–16^. Future studies among individuals with GDM with adequate enrollment of individuals of different BMI categories are needed to clarify whether lipid subclasses are an effect modifier of GDM-associated outcomes that is dependent on maternal adiposity. Studies should also expand investigations to other lipids to further clarify the mechanisms leading to fetal overgrowth due to higher lipids (which lipids, placental transfer, etc.) so novel therapeutic approaches can be developed and tested.

Although not all the studies of insulin profiles made direct statistical comparisons among individuals with GDM, in general, it appears that individuals with GDM who have a defect in insulin sensitivity, (i.e., high insulin resistance), are at increased risk of fetal overgrowth and subsequent delivery of an LGA baby. These studies were from different geographic locations and most diagnosed GDM using IADPSG criteria. There is inadequate data to determine whether isolated insulin secretion defects without concomitant insulin resistance are related to adverse perinatal outcomes. If GDM subtyping based on insulin physiology is to be translated clinically, we need laboratory standardization of insulin (or c-peptide) assays, so we can address the challenge of establishing of a clinical threshold to determine insulin resistance-based GDM subtypes.

Given the established relationship of adiponectin as an insulin sensitizer^111^ and leptin as modulator of food intake and energy expenditure^112^, it is surprising that our review only identified two studies among individuals with GDM that reported associations between adipokines and adverse perinatal outcomes. It is difficult to assess if this reflects a publication bias where null findings have been excluded, or a true lack of research in this area. Indeed, future studies assessing adipose-derived peptides as precision markers among individuals with GDM should also consider additional effect modification by maternal adiposity. This latter point may be particularly relevant as previous studies of adipokines in pregnancy have reported effect modification by maternal BMI^113, 114^.

Notably, other peptide hormones such as glucagon-like 1 peptide, which plays an essential role in glucose homeostasis were absent from the studies reviewed. In addition, no studies that met our inclusion criteria included measures of branched chain amino acids, which have been implicated in diabetes risk and complications both within and outside of pregnancy^115, 116^. Although we recognize that pregnancy cohorts not restricted to GDM only have found associations of amino acids with glucose metabolism and perinatal outcomes^117–121^; However, whether amino acid subclasses or indeed hormonal profiles might be used as potential precision markers among individuals with GDM that identify increased risk of adverse prenatal outcomes has not been adequately studied and future research in this area is needed.

Two candidate gene studies, and two studies of non-coding RNAs suggest that subtyping based on data sources, may identify individuals at higher risk for adverse pregnancy outcomes. The use of omics approaches to subtype individuals with GDM has been limited. Studies performed to date are not only limited in number but have limitations. All have been homogeneous, and in the context of omic studies, the studies have been limited in size.

Moreover, except for Lu et al. who used an array-based approach to identify lncRNAs for prediction of macrosomia, studies to date have used a targeted approach examining either a single or limited number of variants/molecules. Finally, there are no reports integrating different omics technologies for more effective subtyping of individuals with GDM, and none that included metagenomics. There is therefore an important opportunity to integrate omics technologies with other clinical and biochemical measures to better subtype individuals with GDM and identify individuals at high risk for adverse maternal and newborn outcomes.

### Maternal anthropometry/fetal biometry

Most of our included studies evaluated maternal (pre-pregnancy) BMI, and despite low to moderate data quality assessment pertaining to methodology (mostly retrospective single-center hospital-based studies with self-reported or unclear collection/measurement of pre-pregnancy BMI), there was a consistent positive association between pre-pregnancy overweight/obesity with adverse pregnancy and perinatal outcomes, particularly for fetal overgrowth and subsequent LGA or macrosomia. Although assessment of the relative contribution of maternal glycemia versus obesity to adverse pregnancy and perinatal outcomes was beyond the scope of this review; the risks associated with obesity and GDM are additive^61^, which has significant implications given the current obesity epidemic. Importantly, it isn’t obesity per se, but rather the metabolic alterations that accompany obesity that increase risk of adverse perinatal outcomes. This underscores the need to better refine the phenotyping of GDM individuals based on lipids, insulin resistance, and other markers that may participate in fetal overgrowth. While clinically, fetal biometry is not a novel precision marker of overgrowth risk, few research studies have evaluated a combination of early ultra-sonic fetal growth biometry with other metabolic data, in association with, or prediction of, adverse perinatal outcomes. These studies are needed as they may help identify early metabolic biomarker profiles (and therefore targets) of birth size.

Few studies have assessed the association of early GWG among individuals with GDM with regard to outcome beyond GDM diagnosis. From the limited studies reviewed, greater GWG prior to diagnosis may be a risk factor for adverse perinatal outcomes. Taken together, the studies of early GWG and pre-pregnancy obesity confirm the need to target maternal adiposity prior to pregnancy. This is particularly critical in the setting of increasing rates of obesity among individuals of reproductive age.

### Core clinical risk factors, sociocultural and environmental measures

Macrosomia and LGA tended to be more frequent among individuals with GDM who had at least one risk factor. These studies using a composite of risk factors included characteristics such as a prior GDM pregnancy, a family history of diabetes, a prior LGA/macrosomia delivery, or obesity, thus it is unclear if these findings were largely driven by maternal BMI, as reviewed in the prior section. Individually, the presence of factors that increase the risk of GDM in the general population, were not consistently associated with adverse perinatal outcomes. Prior history of macrosomia was associated with risk of higher birth weight (LGA or macrosomia), but not all studies accounted for maternal BMI. As the prevalence of these risk factors is increasing in reproductive age individuals, there may indeed be an overall greater perinatal risk associated with GDM.

Among clinical precision markers, prior history of PCOS in individuals with GDM was associated with preeclampsia. Individuals with both GDM and preeclampsia were at higher risk for SGA compared to individuals with GDM only, in line with known preeclampsia-related risk of fetal growth restriction. Race is a social construct that is recognized to be related to increased risk of GDM (such as Asian, First Nations/Indigenous, Hispanic). The evidence is mixed whether individuals with GDM from these groups are at higher risk of complications; and we note that these racial and ethnic categories and their relationship with outcomes are highly dependent on the overall social context (countries or regions). Future studies with carful collection and consideration of sociocultural influences, such as race, among individuals with GDM are needed.

### Limitations

It is worthwhile to note that half of the studies were considered low quality, which impacts the quality of conclusions drawn from the data. Studies were often rated as low quality due to unclear reporting of methods or presenting unadjusted estimates, which can be a source of bias in observational studies. These impacts on quality are because most studies provided the data relevant to the current review in a sub-analysis only, and therefore thorough confounder consideration or reporting of data collection methods for the variables of interest to the current study were frequently not included in the written manuscripts.

### Future directions and overall conclusions

Our systematic review has identified several major areas for further research. First, there is a need for studies among individuals with GDM that collect biospecimens longitudinally that can be used for comprehensive measurement of multi-omics markers. Such data are pivotal for an in-depth and systematic understanding of precision biomarkers for GDM and subsequent pregnancy and perinatal outcomes. Related to this is a need for standardization of laboratory analysis for biomarker assessment; mechanistic studies restricted to GDM cases to understand differences in pathophysiology that contribute to heterogeneity in outcomes; and the inclusion of larger and more diverse populations. Second, studies with measurement of genetics and multi-omics that integrate clinical and sociocultural data are needed and could provide insight into the determinants and causal pathways of heterogeneity within GDM pregnancies. This latter need may require applying approaches often used in systems biology or in the aggregation and analysis of large datasets from different sources. Lastly, there were a limited number of studies measuring early pregnancy sociocultural factors such as dietary intake, deprivation, environmental influences, and other lifestyle behaviors, which impact perinatal outcomes and may explain variation among individuals with GDM.

There are currently limited systematic reviews of precision markers related to GDM diagnosis and adverse perinatal outcomes. Findings from the current study demonstrate that individuals with adiposity who develop GDM are at a substantially higher risk of LGA or macrosomia than those with GDM and lower adiposity, highlighting the need for innovative prevention and intervention strategies. An overarching theme was a lack of studies integrating data across all domains of precision markers. Advances in computing and the promotion of cross-disciplinary team science may be one approach for addressing these gaps and future directions. Given the global and transgenerational burden of GDM, and the increasing prevalence of GDM risk factors, identification of precision markers for GDM diagnosis will clarify whether precision medicine in pregnancy can result in a refined detection of GDM and its subtypes.

The risk of bias and overall quality of each study was assessed independently or in duplicate using the Joanna Briggs Institute (JBI) Critical Appraisal Tool for cohort studies, which was modified specifically for the objectives of the current systematic review. For each question, a reviewer could indicate “not applicable: 0”, “yes: 1”, “unclear: 2”, “no: 3”. An answer of “yes” indicates less risk of bias and greater quality, and answer of “no” indicates a higher risk of bias and lower quality

## Data Availability

All data included in this systematic review are publicly available.

