## Supplementary material for "Refining the diagnosis of Gestational Diabetes Mellitus: A systematic review to inform efforts in precision medicine"

Supplementary Table 1: Characteristics and narrative of studies included as biochemical, genetics, omic..…..…………..Page 1-2.

Supplementary Table 2: Characteristics and narrative of studies included as maternal/fetal anthropometry..…………..…Page 3-5.

Supplementary Table 3: Characteristics and narrative of studies included as clinical, sociocultural, environmental……..Page 6-7.

Supplementary Figure 1: Quality assessment of studies included as biochemical, genetics, omic..………………………Page 8.

Supplementary Figure 2: Quality assessment of studies included as maternal/fetal anthropometry..…………………..…Page 9.

Supplementary Figure 3: Quality assessment of studies included as clinical, sociocultural, environmental..…………..…Page 10.

Supplementary Material 1: Search strategy……………………………………………………………………………...Page 11-22.

Supplementary Material 2: Modified Joanna Briggs Institute critical appraisal checklist for cohort studies..………..……Page 23-25.

**Supplementary Table 1: Characteristics and brief narrative of studies included as biochemical, genetics, 'omic**

| Author, yr. | Enrollment yrs. | Country | Total GDM cases | GDM diagnostic criteria | Age | BMI a | nulliparous (%) |
|---|---|---|---|---|---|---|---|
| **Lipids** | | | | | | | |
| Barden 2004 | NR | Australia | 184 | ADIPS | NR | NR | NR |
| Gorban de Lapertosa 2020 | 2017-2019 | Argentina | 1088 | NR | NR | 28% ¥ | NR |
| Grotenfelt 2019 | 2008-2014 | Finland | 164 | IADPSG | NR | 100% ¥ | NR |
| Hashemipour 2018 | 2015-2016 | Iran | 305 | WHO/IADPSG | NR | 19.3% ¥ | NR |
| HerreraMartínez 2018 | 2013-2015 | Spain | 250 | Spanish Group of Diabetes and Pregnancy | 33.08 | 27.67 | 0.435 |
| Knopp 1992 | 1985 - 1986 | USA | 96 | CC | NR | 23.8 | NR |
| Krstevska 2009 | 2006-2009 | Macedonia | 200 | CC | NR | 26.64 | NR |
| Olmos 2014 | 2009-2013 | Chile | 279 | 2-hour glucose >= 140 mg/dl (7.77 mmol/l) | NR | 16.5% ¥ | NR |
| Rao 2021 | 2016-2018 | China | 565 | IADPSG | 31.9 | 21.9 | NR |
| Simeonova-Krstevska 2014 | NR | Macedonia | 200 | IADPSG | 31.3 | 26.9 | NR |
| Son 2010 | 2000-2008 | Korea | 104 | CC | 32.7 | 23.2 | NR |
| Sun 2020 a | 2014 - 2016 | China | 2647 | IADPSG | 32.31 | NR | NR |
| Xiao 2020 | 2016-2018 | China | 248 | IADPSG | NR | 43.1% ¥ | NR |
| Zawiejska 2008 | 1993-2005 | Poland | 357 | WHO/IADPSG | NR | 24.2 | NR |
| Zou 2021 | 2019-2020 | China | 783 | IADPSG | NR | NR | NR |
| **Insulin profiles** | | | | | | | |
| Benhalima 2019 a | NR | Belgium | 228 | WHO/IADPSG | NR | NR | NR |
| Bomba-Opon 2010 | NR | Poland | 121 | Polish Gynaecological Society | 31.1 | 24.03 | NR |
| Gibbons 2021 | NR | Multinational b | 1026 | IADPSG | 29.4 | 26.9 | 0.473 |
| Immanuel 2021 | 2012-2014 | Multinational c | 236 | IADPSG | NR | 34 | NR |
| Knopp 1992 | 1985 - 1986 | USA | 96 | CC | NR | 23.8 | NR |
| Li 2018 | 2010-2012 | China | 923 | IADPSG | 29.8 | 10% ¥ | 0.956 |
| Lin 2021 | 2018-2019 | China | 710 | IADPSG | 33 | 7% ¥ | 0.587 |
| Liu 2018 | 2015-2016 | China | 206 | IADPSG | 29 | 22 | 0.66 |
| Madsen 2021 | 199-2006 | Multinational a | 1090 | WHO/IADPSG | 29.4 | 27 | 0.469 |
| Sun 2020 a | 2014 - 2016 | China | 2647 | IADPSG | 32.31 | NR | NR |
| Wang 2021 b | 2019-2020 | China | 299 | IADPSG | NR | NR | NR |
| Zhang 2020 | 2015-2018 | China | 737 | IADPSG | 31.1 | NR | NR |
| **Adipokines** | | | | | | | |
| Bomba-Opon 2010 | NR | Poland | 121 | Polish Gynaecological Society | 31.1 | 24.03 | NR |

| | | | | | | | |
|---|---|---|---|---|---|---|---|
| Park 2013 | 2006 - 2009 | Korea | 215 | CC | NR | 24.7 | NR |
| **Genetics, omics** | | | | | | | |
| Bo 2015 | 2009-2012 | Italy | 200 | IADPSG | NR | NR | NR |
| Han 2015 | 2011 - 2012 | China | 128 | IADPSG | 27.9 | NR | NR |
| Lu 2018 | 2011-2016 | China | 600 | IADPSG | NR | NR | NR |
| Sun 2020 | 2017-2018 | China | 64 | IADPSG | 34.3 | 23.8 | 0.76 |
| Zhu 2021 | 2019-2020 | China | 70 | IADPSG | 27.8 | 21.4 | 0.68 |

Footnotes:
a BMI is reported as mean/median or else as % : ¥ % of women with GDM with a BMI ≥ 30.0 $^{kg/m2}$ ; § % of women with a GDM with a BMI ≥ 25.0 $^{kg/m2}$

Multinational a: Bellflower, CA, USA; Cleveland, OH, USA; Brisbane, QLD, Australia; Newcastle, NSW, Australia; Hong Kong, China
Multinational b: HAPO (Cleveland, Bellflower, Brisbane, Newcastle, Hong Kong)
Multinational c: Spain, UK, Austria, Belgium, Denmark, Italy, Ireland, Poland, Netherlands

**Supplementary Table 2: Characteristics and brief narrative of studies included as maternal anthropometry/fetal biometry**

| Author, yr. | Enrollment yrs. | Country | Total GDM cases | Diagnostic criteria | Age Mean/median | BMI a | nulliparous (%) |
|---|---|---|---|---|---|---|---|
| **Maternal Body Mass Index** | | | | | | | |
| Alfadhli 2021 | 2014-2015 | Saudi Arabia | 266 | WHO/IADPSG | 30.5 | 29.3 | NR |
| Antoniou 2020 | 2012-2017 | Switzerland | 189 | IADPSG | 32.9 | 26.6 | NR |
| Barden 2004 | NR | Australia | 184 | ADIPS | NR | NR | NR |
| Barnes 2013 | 1992-2009 | Australia | 1695 | ADIPS | NR | 26.1 | NR |
| Barquiel 2014 | 1987 - 2008 | Spain | 2037 | NDDG | NR | 24.7 | NR |
| Barquiel 2016 | 1987 - 2008 | Spain | 2037 | NDDG | 33 | 24.7 | NR |
| Barquiel 2018 | 1986 - 2015 | Spain | 3284 | NDDG | NR | 11.1% ¥ | NR |
| Ben-Haroush 2009 | 1993-2004 | Israel | 233 | CC | NR | NR | NR |
| Blickstein 2018 | 2003-2012 | Slovenia | 6229 | CC | NR | 69.8% ¥ | NR |
| Catalano 2012 | 2000-2006 | Multinational [a] | 3746 | WHO/IADPSG | 29.2 | 27.7 | 47.5% |
| Chee 2020 | 2004-2015 | Australia | 1064 | ADIPS (1998 ) | NR | NR | 36.7% |
| Collins 2018 | 2014-2016 | Australia | 410 | IADPSG | NR | 25.4% ¥ | NR |
| Cosson 2016 | 2002-2010 | France | 2097 | CNGOF | 29.7 | 24.6 | 41.8% |
| Cremona 2020 | 2016 | Ireland | 303 | IADPSG | 33.6 | 27.8 | 19.8% |
| dePaulaBertoli 2020 | 2010 - 2018 | Brazil | 442 | NR | NR | 58.8% ¥ | NR |
| Ducarme 2018 | 2014-2015 | France | 200 | WHO/IADPSG | 31 | 28 | 36.0% |
| Filardi 2018 | 2014-2016 | Italy | 183 | Other | 33.8 | 27.9% ¥ | 37.8% |
| Fonseca 2021 | 2011-2017 | Portugal | 1085 | IADPSG | 32.9 | 26.5 | 86.1% |
| Fuka 2020 | 2013-2014 | Fiji | 255 | Modified IADPSG | 30.7 | 33.2 | 13.3% |
| García-Patterson 2004 | 1986-2022 | Spain | 2060 | 3rd Work-shop-Conference on GDM criteria | NR | NR | NR |
| García-Patterson 2012 | 1986-2006 | Spain | 2092 | NDDG | NR | NR | NR |
| Gascho 2017 | NR | Brazil | 392 | IADPSG | NR | NR | NR |
| Gorban de Lapertosa 2020 | 2017-2019 | Argentina | 1088 | NR | NR | 28% ¥ | NR |
| Grotenfelt 2019 | 2008-2014 | Finland | 164 | IADPSG | NR | 100% ¥ | NR |
| Hagiwara 2018 | 2011-2016 | Japan | 675 | IADPSG | NR | 27.7% § | 44.0% |
| Hardy 1999 | 1993-1994 | USA | 213 | 3-hr OGTT [thresholds not provided] | NR | NR | NR |
| Hashemipour 2018 | 2015-2016 | Iran | 305 | WHO/IADPSG | NR | 19.3% ¥ | NR |
| Hernandez-Rivas 2013 | 2004-2011 | Spain | 456 | NDDG | NR | NR | NR |

| | | | | | | | |
|---|---|---|---|---|---|---|---|
| Hildén 2016 | 1998-2012 | Sweden | NR | ICD-10 codes | NR | 30.4% § | NR |
| Hod 1996 | 1986-1990 | Israel | 470 | ADA (1979 to 1985) & ACOG (1986) | NR | NR | NR |
| Huet 2018 | 2012-2014 | France | 808 | CNGOF (2010) | NR | 32.1% ¥ | NR |
| Ijäs 2019 | 2009 | Finland | 5680 | Finnish national guidelines (2008) | NR | 22% ¥ | NR |
| Krstevska 2009 | 2006-2009 | Macedonia | 200 | CC | NR | 26.64 | NR |
| Langer 2016 | 1990-1999 | USA | 555 | CC | NR | 28.2% ¥ | 21.5% |
| Leng 2015 | 2009-2011 | China | 1263 | Other | NR | 8.3% ¥ | NR |
| Li 2021 | 2018-2020 | China | 16829 | IADPSG | NR | 3.9% ¥ | 48.5% |
| Martin 2015 | 2008-2011 | Australia | 115 | South Australian state-wide perinatal practice guidelines | 29.6 | 56.8% ¥ | 40.2% |
| Masalin 2019 | 2009-2015 | Finland | NR | Finnish Current Care Guidelines | NR | NR | 80.1% |
| Matta-Coelho 2019 | 2011 - 2015 | Portugal | 10443 | IADPSG | 33.2 | 26 | NR |
| Much 2015 | 1998 - 2010 | Germany | 856 | WHO/IADPSG | 32.8 | 29.5 | NR |
| Mustaniemi 2021 | 2009-2012 | Finland | 1055 | Finnish Current Care guidelines | NR | 28.1 | 43.1% |
| Nobumoto 2015 | 2001 - 2011 | Japan | 446 | Older IADPSG | 30.26 | 21.1 | NR |
| Nunes 2020 | 2017-2018 | Portugal | 301 | Medical records | 33.37 | 25.6% ¥ | NR |
| Olmos 2012 | 1998-2009 | Chile | 251 | WHO 1999 | NR | 50.2% § | NR |
| Olmos 2014 | 2009-2013 | Chile | 279 | 2-hour glucose >= 140 mg/dl (7.77 mmol/l) | NR | 16.5% ¥ | NR |
| Ouzounian 2011 | 2000 - 2006 | USA | 1502 | NR | 32.1 | 28.7 | NR |
| Pezzarossa 1996 | NR | Italy | 60 | NDDG | 28 | 25.4 | NR |
| Phaloprakarn 2009 | 2003-2008 | Thailand | 813 | CC | NR | NR | NR |
| Philipson 1985 | 1979 - 1983 | USA | 158 | NR | NR | 39.2% ¥ | NR |
| Pintaudi 2018 | 2012-2015 | Italy | 2736 | IADPSG | 36.6 | 24.8 | 45.3% |
| Quaresima 2020 | 2018-2020 | Italy | 219 | IADPSG | 31.7 | 25.8 | 62.50% |
| Schaefer-Graf 2011 | 2001-2007 | Germany | 1914 | CC | NR | NR | NR |
| Scifres 2015 | 2009-2012 | USA | 1344 | CC | NR | 47.4% ¥ | NR |
| Simeonova-Krstevska 2014 | NR | Macedonia | 200 | IADPSG | 31.3 | 26.9 | NR |
| Son 2010 | 2000-2008 | Korea | 104 | CC | 32.7 | 23.2 | NR |
| Sun 2014 | 2010-2012 | China | 1418 | IADPSG | NR | 25.6% § | NR |
| Tavares 2019 | 2015-2017 | Brazil | 116 | IADPSG | 32.7 | 43% ¥ | 75.0% |

| | | | | | | | |
|---|---|---|---|---|---|---|---|
| Thevarajah 2019 | 2013-2015 | Australia | 749 | WHO/IADPSG; ADIPS guidelines | NR | 31.7% ¥ | 27.8% |
| Usami 2020 | 2003-2009 | Japan | 1481 | Former SOG | NR | NR | NR |
| Wahabi 2014 | 2011-2012 | Saudi Arabia | 415 | CC | 29.38 | 30.25 | NR |
| Wang 2015 | 2012-2013 | China | 587 | WHO/IADPSG | 30.2 | 22.66 | 93.2% |
| Wang 2018 | 2014 | China | 601 | IADPSG | 30.5 | 21.7 | 83.9% |
| Weschenfelder 2021 | 2012-2016 | Germany | 614 | WHO/IADPSG | 31 | 26.6 | NR |
| Yogev 2004 b | 1994-1999 | USA | 1664 | CC | 28.7 | 27.7 | NR |
| Yogev 2008 | 1990-1999 | USA | 1319 | CC | NR | 100% ¥ | NR |
| Yue 2022 | 2016-2018 | Vietnam | 908 | WHO/IADPSG | NR | 23.5 | 53.5% |
| Yuen 2021 | 2007-2008 | New Zealand & Australia | 7518 | Waikato NZSSD 1995, Waikato NZMOH 2014, ADIPS 1991, IADPSG | NR | NR | NR |
| Zawiejska 2008 | 1993-2005 | Poland | 357 | WHO/IADPSG | NR | 24.2 | NR |
| Zou 2021 | 2019-2020 | China | 783 | IADPSG | NR | NR | NR |
| **Early Gestational Weight Gain** | | | | | | | |
| Aiken 2019 | 2014-2017 | UK | 129 | IADPSG with a higher fasting threshold ≥5.3 mmol/l | 34.3 | 30.9 | 36.6% |
| Barnes 2013 | 1992-2009 | Australia | 1695 | ADIPS | NR | 26.1 | NR |
| Horosz 2013 | 2005-2011 | Poland | 675 | Polish Gynaecological Society | 31.5 | 25.3 | 51.0% |
| Shi 2021 | 2010-2020 | China | 1606 | IADPSG | 32.75 | 22.72 | 58.9% |
| **Fetal Biometry** | | | | | | | |
| Antoniou 2020 | 2012-2017 | Switzerland | 189 | IADPSG | 32.9 | 26.6 | NR |
| Bomba-Opon 2010 | NR | Poland | 121 | Polish Gynaecological Society | 31.1 | 24.03 | NR |
| Chee 2020 | 2004-2015 | Australia | 1064 | ADIPS (1998 ) | NR | NR | 36.7% |
| Gascho 2017 | NR | Brazil | 392 | IADPSG | NR | NR | NR |
| Lee 2014 | 2006-2013 | South Korea | 243 | CC | NR | 23.5 | 57.0% |
| Leung 2004 | 2002 | China | 138 | Other | NR | NR | NR |
| Schaefer-Graf 2011 | 2001-2007 | Germany | 1914 | CC | NR | NR | NR |
| Simpson 2018 | 2012-2015 | USA | 413 | IADPSG | 30.1 | 32.6 | 20.8% |
| Zou 2021 | 2019-2020 | China | 783 | IADPSG | 0 | NR | NR |

Footnotes: a BMI is reported as mean/median or else as % : ¥ % of women with GDM with a BMI ≥ 30.0 $^{kg/m2}$ ; § % of women with a GDM with a BMI ≥ 25.0 $^{kg/m2}$
Multinational a: USA, Canada, West Indies, UK, Israel, Singapore, Thailand, China, Australia

**Supplementary Table 3: Characteristics and brief narrative of studies included as clinical, sociocultural, environmental**

| Author, yr. | Enrollment yrs. | Country | Total GDM cases | GDM diagnostic criteria | Age | BMI a | nulliparous (%) |
|---|---|---|---|---|---|---|---|
| **Compositive of multiple clinical/sociocultural risk factors** | | | | | | | |
| Benhalima 2019 b | 2014-2017 | Belgium | 231 | WHO/IADPSG | NR | 10.2% ¥ | NR |
| Cosson 2013 | 2002-2010 | France | 2710 | WHO/IADPSG CGNOF | 29.7 | 24.1 | NR |
| Hammoud 2013 | 2006-2009 | Netherlands | 249 | NR | 33.1 | 27.9 | 27.7% |
| Matta-Coelho 2019 | 2011 - 2015 | Portugal | 10443 | IADPSG | 33.2 | 26 | NR |
| Quaresima 2020 | 2018-2020 | Italy | 219 | IADPSG | 31.7 | 25.8 | 62.5% |
| Weeks 1994 | 1990-1992 | USA | 106 | NDDG | 30.4 | 7% ¥ | NR |
| **Unique clinical/sociocultural modifiers** | | | | | | | |
| Kouhkan 2018 | 2014-2017 | Iran | 287 | IADPSG | NR | NR | NR |
| Li 2010 | 2006-2009 | China | 104 | CC | NR | NR | NR |
| Liu 2020 | 2014 - 2018 | China | 314 | IADPSG | 32.1 | 22.9 | 100.0% |
| Meek 2020 | 2004-2008 | UK | 985 | WHO 1999 | 32.6 | 22% ¥ | 39.4% |
| Szymanska 2011 | NR | Poland | 173 | NR | NR | NR | 91.0% |
| Tundidor 2012 | 1981-2007 | Spain | 2299 | NR | 0 | 23.31 | 36.7% |
| Yogev 2004 a | 1993-1999 | USA | 1813 | CC | NR | NR | NR |
| Zhang 2017 | 2005-2009 | China | 1263 | WHO/IADPSG | 0 | 22.9 | NR |
| **Psychosocial risk factors** | | | | | | | |
| Cosson 2015 | 2009-2012 | France | 994 | IADPSG; CGNOF | 33.3 | 27.8 | 26.8% |
| Lee 2020 | 2018 | Malaysia | 418 | NR | 32.4 | NR | NR |
| Packer 2021 | 2007 - 2011 | USA | 170572 | NR | NR | NR | NR |
| Schmidt 2019 | NR | Netherlands | 100 | NR | 32.5 | 26.7 | 39.0% |
| **Individual clinical or sociocultural risk factors** | | | | | | | |
| Alshammari 2010 | 1999-2006 | Canada | 171 | O'Sullivan or CDA | NR | NR | 50.3% |
| Anyaegbunam 1995 | 1990-1992 | USA | 418 | NDDG | NR | NR | NR |
| Berggren 2012 | NR | USA | 768 | CC | NR | NR | NR |
| Chee 2020 | 2004-2015 | Australia | 1064 | ADIPS (1998 ) | NR | NR | 36.7% |
| Chen 2019 | 1996-2010 | Canada | 12110 | Canadian | NR | NR | NR |
| Contreras 2010 | 2003-2006 | USA | 915 | CC | NR | 26% ¥ | 31.5% |
| Ding 2018 | 2015 - 2017 | China | 3221 | WHO/IADPSG | 32.7 | NR | NR |
| Ducarme 2018 | 2014-2015 | France | 200 | WHO/IADPSG | 31 | 28 | 36.0% |
| Dyck 2020 | 1980-2013 | Canada | 10514 | ICD-9 codes | NR | NR | 32.0% |

| | | | | | | | |
|---|---|---|---|---|---|---|---|
| Esakoff 2011 | 2001-2004 | USA | 26411 | NR | NR | NR | NR |
| Fadl 2012 | 1998-2007 | Sweden | 8560 | DPSG | NR | NR | NR |
| Filardi 2018 | 2014-2016 | Italy | 183 | Other | 33.8 | 27.9% ¥ | 37.8% |
| Fraser 1994 | 1987-1988 | Israel | 442 | NDDG | NR | NR | NR |
| Gorban de Lapertosa 2020 | 2017-2019 | Argentina | 1088 | NR | 0 | 28% ¥ | NR |
| Hernandez-Rivas 2013 | 2004-2011 | Spain | 456 | NDDG | NR | NR | NR |
| Kwong 2019 | 2009-2013 | Canada | 537 | CDA & SOGC (2008 ) | 34.1 | NR | NR |
| Lamminpää 2016 | 2004 - 2008 | Finland | 27154 | ICD-10 codes and text searching | NR | NR | NR |
| Makgoba 2012 | 1988-2000 | UK | 1113 | Varied across sites | NR | NR | NR |
| Manoharan 2020 | 2015-2019 | Australia | 1545 | IADPSG; ADIPS | NR | NR | NR |
| Mocarski 2012 | 2001-2006 | USA | 19416 | NR | NR | 13.7% ¥ | NR |
| Nunes 2020 | 2017-2018 | Portugal | 301 | Medical records | 33.37 | 25.6% ¥ | NR |
| Pintaudi 2018 | 2012-2015 | Italy | 2736 | IADPSG | 36.6 | 24.8 | 45.3% |
| Quaresima 2020 | 2018-2020 | Italy | 219 | IADPSG | 31.7 | 25.8 | 62.5% |
| Rao 2021 | 2016-2018 | China | 565 | IADPSG | 31.9 | 21.9 | NR |
| Scime 2020 | 2014-2017 | Canada | 11114 | NR | 0 | NR | NR |
| Thevarajah 2019 | 2013-2015 | Australia | 749 | WHO/IADPSG; ADIPS guidelines | 0 | 31.7% ¥ | 27.8% |
| Tsai 2013 | 2009 - 2011 | Hawaii | 5925 | Self-report | NR | NR | NR |
| Wan 2019 | 2010-2013 | Australia | 1579 | ADIPS (1991) | NR | NR | NR |
| Wang 2021 a | 2012-2013 | China | 1229 | IADPSG | 0 | 1.9% § | 81.8% |
| Yuen 2021 | 2007-2008 | New Zealand & Australia | 7518 | Waikato NZSSD 1995, Waikato NZMOH 2014, ADIPS 1991, IADPSG | NR | NR | NR |
| Zawiejska 2008 | 1993-2005 | Poland | 357 | WHO/IADPSG | 0 | 24.2 | NR |

Footnotes: a BMI is reported as mean/median or else as % : ¥ % of women with GDM with a BMI ≥ 30.0 $^{kg/m2}$ ; § % of women with a GDM with a BMI ≥ 25.0 $^{kg/m2}$

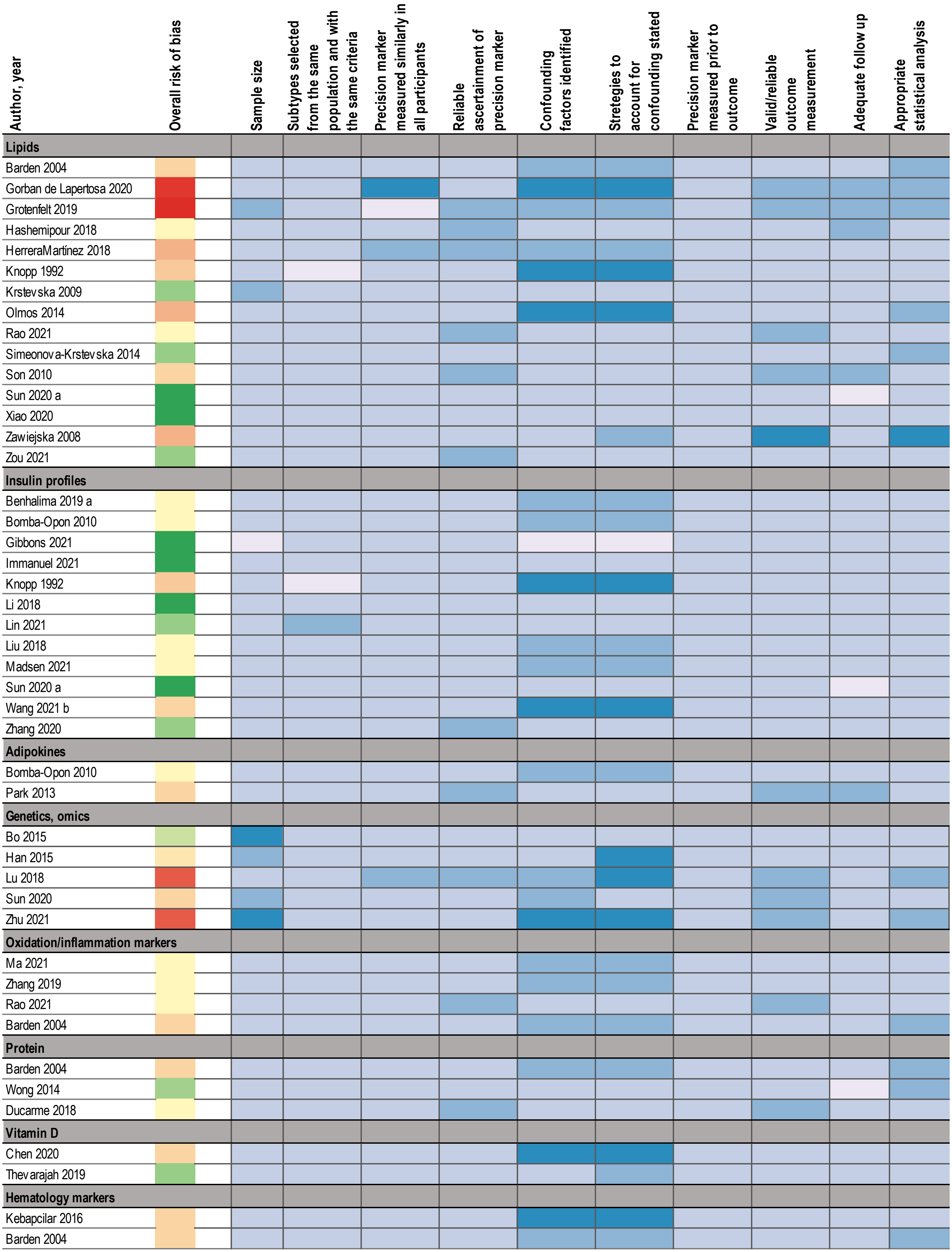


**Supplementary Figure 1: Quality assessment of studies included as biochemical, genetics, omic**

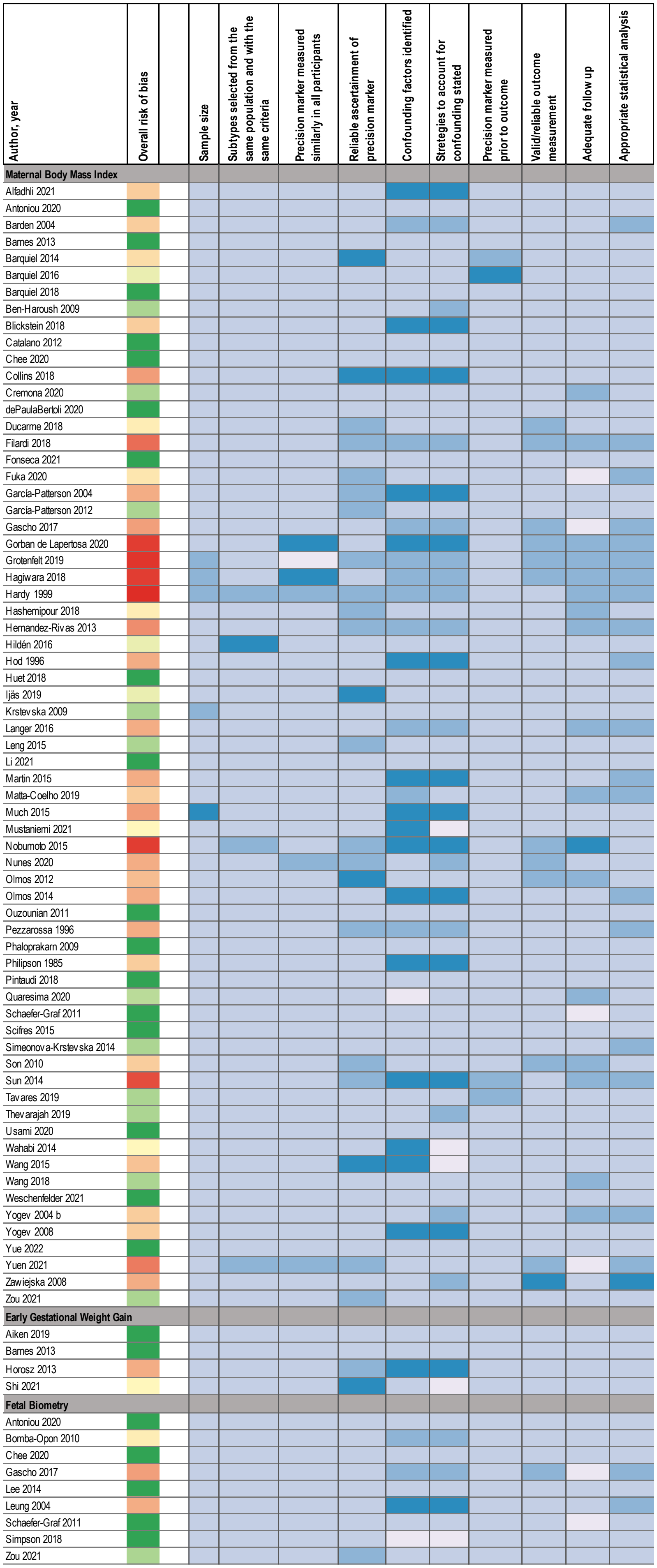


**Supplementary Figure 2: Quality assessment of studies included as maternal/fetal anthropometry**

| Author, year | Overall risk of bias | | Sample size | Subtypes selected from the same population and with the same criteria | Precision marker measured similarly in all participants | Reliable ascertainment of precision marker | Confounding factors identified | Stretegies to account for confounding stated | Precision marker measured prior to outcome | Valid/reliable outcome measurement | Adequate follow up | Appropriate statistical analysis |
|---|---|---|---|---|---|---|---|---|---|---|---|---|
| **Compositive of multiple clinical or sociocultural risk factors** | | | | | | | | | | | | |
| Benhalima 2019 b | | | | | | | | | | | | |
| Cosson 2013 | | | | | | | | | | | | |
| Hammoud 2013 | | | | | | | | | | | | |
| Matta-Coelho 2019 | | | | | | | | | | | | |
| Quaresima 2020 | | | | | | | | | | | | |
| Weeks 1994 | | | | | | | | | | | | |
| **Novel/unique clinical or sociocultural modifiers** | | | | | | | | | | | | |
| Kouhkan 2018 | | | | | | | | | | | | |
| Li 2010 | | | | | | | | | | | | |
| Liu 2020 | | | | | | | | | | | | |
| Meek 2020 | | | | | | | | | | | | |
| Szymanska 2011 | | | | | | | | | | | | |
| Tundidor 2012 | | | | | | | | | | | | |
| Yogev 2004 a | | | | | | | | | | | | |
| Zhang 2017 | | | | | | | | | | | | |
| **Psychosocial risk factors** | | | | | | | | | | | | |
| Cosson 2015 | | | | | | | | | | | | |
| Lee 2020 | | | | | | | | | | | | |
| Packer 2021 | | | | | | | | | | | | |
| Schmidt 2019 | | | | | | | | | | | | |
| **Individual clinical or sociocultural risk factors** | | | | | | | | | | | | |
| Alshammari 2010 | | | | | | | | | | | | |
| Anyaegbunam 1995 | | | | | | | | | | | | |
| Berggren 2012 | | | | | | | | | | | | |
| Chee 2020 | | | | | | | | | | | | |
| Chen 2019 | | | | | | | | | | | | |
| Contreras 2010 | | | | | | | | | | | | |
| Ding 2018 | | | | | | | | | | | | |
| Ducarme 2018 | | | | | | | | | | | | |
| Dyck 2020 | | | | | | | | | | | | |
| Esakoff 2011 | | | | | | | | | | | | |
| Fadl 2012 | | | | | | | | | | | | |
| Filardi 2018 | | | | | | | | | | | | |
| Fraser 1994 | | | | | | | | | | | | |
| Gorban de Lapertosa 2020 | | | | | | | | | | | | |
| Hernandez-Rivas 2013 | | | | | | | | | | | | |
| Kwong 2019 | | | | | | | | | | | | |
| Lamminpää 2016 | | | | | | | | | | | | |
| Makgoba 2012 | | | | | | | | | | | | |
| Manoharan 2020 | | | | | | | | | | | | |
| Mocarski 2012 | | | | | | | | | | | | |
| Nunes 2020 | | | | | | | | | | | | |
| Pintaudi 2018 | | | | | | | | | | | | |
| Quaresima 2020 | | | | | | | | | | | | |
| Rao 2021 | | | | | | | | | | | | |
| Scime 2020 | | | | | | | | | | | | |
| Thevarajah 2019 | | | | | | | | | | | | |
| Tsai 2013 | | | | | | | | | | | | |
| Wan 2019 | | | | | | | | | | | | |
| Wang 2021 a | | | | | | | | | | | | |
| Yuen 2021 | | | | | | | | | | | | |
| Zawiejska 2008 | | | | | | | | | | | | |

**Supplementary Figure 3: Quality assessment of studies included as clinical, sociocultural, environmental**

**Supplementary Material 1: Search strategy**

### GDM Precision diagnosis

#### Search strategy Embase (Elsevier)

Date of search: 211005, updated 211202, 220210, 220221, 220223, 220302, 220321

**Research question 1**

#1 'pregnancy diabetes mellitus'/exp

43097 records

#2 'gestational diabetes':ab,ti OR gdm:ab,ti OR 'pregnancy induced diabetes':ab,ti OR 'pregnancy-induced diabetes':ab,ti

29110 records

#3 #1 OR #2

46421 records

**1A How can the diagnosis of GDM be refined?**

**BMI, biomarkers, phenotypes age, ethnic*, sociocultural, behavioral factors, diet, exercise/physical activity block**

#4 'phenotype'/exp OR 'genetic heterogeneity'/exp OR 'timing'/exp OR 'severity'/exp OR 'insulin resistance'/exp OR 'insulin sensitivity'/exp OR 'insulin release'/exp

1267188 records

#5 phenotyp*:ab,ti OR heterogeneity:ab,ti OR timing:ab,ti OR severity:ab,ti OR 'clinical characteristics':ab,ti OR 'insulin resistance':ab,ti OR 'insulin sensitivity':ab,ti OR 'secretory defect':ab,ti

2239071 records

#6 #4 OR #5

2813362 records

**Biomarkers**

#7 'biological marker'/exp OR 'biological marker*':ab,ti OR biomarker*:ab,ti

610910 records

**Lipids**

#8 'triacylglycerol'/exp OR 'very low density lipoprotein'/exp OR 'cholesterol'/exp OR 'lipoprotein'/exp OR 'high density lipoprotein'/exp OR 'apolipoprotein'/exp OR 'hyperlipidemia'/exp OR 'dyslipidemia'/exp OR 'fatty acid'/exp OR 'glycolysated protein' OR 'lipid'/exp OR lipid*:ab,ti OR triacylglycerol:ab,ti OR triglyceride:ab,ti OR vldl:ab,ti OR 'very low density lipoprotein':ab,ti OR cholesterol:ab,ti OR lipoprotein*:ab,ti OR hdl:ab,ti OR 'high density lipoprotein':ab,ti OR hyperlipidemia:ab,ti OR apolipoprotein*:ab,ti OR 'non hdl':ab,ti OR lipidemia*:ab,ti OR lipemia*:ab,ti OR lipemic:ab,ti OR dyslipidemia:ab,ti OR 'fatty acid*':ab,ti OR 'glycolysated protein*':ab,ti

2247395 records

**Amino acids**

#9 'branched chain amino acid'/exp OR 'creatine'/exp OR 'leucine'/exp OR 'isoleucine'/exp OR 'valine'/exp OR 'alanine'/exp OR 'glutamine'/exp OR 'glutamic acid'/exp OR 'betaine'/exp OR 'aspartic acid'/exp OR 'tyrosine'/exp OR 'glutathione'/exp OR 'serine'/exp OR 'threonine'/exp OR 'histidine'/exp OR 'tryptophan'/exp OR 'glycine'/exp OR 'amino acid'/exp OR 'proteomics'/exp OR proteomic*:ab,ti OR 'branched chain amino acids':ab,ti OR bcaa:ab,ti OR creatine:ab,ti OR leucine:ab,ti OR isoleucine:ab,ti OR valine:ab,ti OR alanine:ab,ti OR glutamine:ab,ti OR glutamate:ab,ti OR betaine:ab,ti OR aspartate:ab,ti OR tyrosine:ab,ti OR glutathione:ab,ti OR serine:ab,ti OR threonine:ab,ti OR histidine:ab,ti OR tryptophan:ab,ti OR glycine:ab,ti OR 'amino acids':ab,ti

2748236 records

**Inflammation**

#10 'cytokine'/exp OR 'c reactive protein'/exp OR 'high sensitivity c reactive protein'/exp OR 'tumor necrosis factor'/exp OR 'mcp1 protein'/exp OR 'chemokine'/exp OR 'inflammation'/exp OR inflammation:ab,ti OR cytokine*:ab,ti OR interleukin*:ab,ti OR il*:ab,ti OR crp:ab,ti OR 'high-sensitivity crp':ab,ti OR 'high sensitivity crp':ab,ti OR 'c-reactive protein':ab,ti OR 'hs crp':ab,ti OR 'tnf alpha':ab,ti OR 'tumor necrosis factor alpha':ab,ti OR 'mcp 1':ab,ti OR 'monocyte chemoattractant protein 1':ab,ti OR chemokine:ab,ti

6650634 records.

**Adipokines or adipocytokines**

#11 'adiponectin'/exp OR 'leptin'/exp OR 'retinol binding protein 4'/exp OR 'adipocytokine'/exp OR adiponectin:ab,ti OR leptin:ab,ti OR rbp4:ab,ti OR 'retinol binding protein 4':ab,ti OR adipokine*:ab,ti OR adipocytokine*:ab,ti

101544 records.

**Hormones**

#12 'sex hormone binding globulin'/exp OR 'estrogen'/exp OR 'hydrocortisone'/exp OR 'prolactin'/exp OR 'progesterone'/exp OR 'somatomedin'/exp OR 'hormone'/exp OR 'hormone* sex hormone-binding globulin':ab,ti OR shbg:ab,ti OR estrogen:ab,ti OR oestrogen:ab,ti OR cortisol:ab,ti OR prolactin:ab,ti OR progesterone:ab,ti OR 'insulin-like growth factor':ab,ti OR igf:ab,ti

799750 records.

**Placenta-derived**

#13 'human placenta lactogen'/exp OR 'placental growth hormone'/exp OR 'placenta derived':ab,ti OR 'human placental lactogen':ab,ti OR hpl:ab,ti OR 'human chorionic somatomammotropin':ab,ti OR hsc:ab,ti OR 'pregnancy-associate plasma protein a':ab,ti OR 'papp a':ab,ti OR 'placental growth hormone':ab,ti OR pgh:ab,ti

34650 records

**Thyroid**

#14 'thyroid gland'/exp OR 'thyroglobulin antibody'/exp OR 'thyroid peroxidase antibody'/exp OR 'free thyroxine index'/exp OR 'free liothyronine index'/exp OR 'thyrotropin'/exp OR thyroid*:ab,ti OR 'thyroid stimulating hormone':ab,ti OR thyrotropin:ab,ti OR tsh:ab,ti OR 'thyroglobulin antibody':ab,ti OR 'thyroid peroxidase antibody':ab,ti OR 'free t4':ab,ti OR 'free t3':ab,ti

319602 records.

**Metabolomics**

#15 'butyric acid'/exp OR 'acetylcarnitine'/exp OR 'acylcarnitine'/exp OR 'carnitine'/exp OR 'lactic acid'/exp OR 'metabolomics'/exp OR metabolomics:ab,ti OR butyrate:ab,ti OR 'butyric acid':ab,ti OR acetylcarnitine:ab,ti OR acylcarnitine:ab,ti OR carnitine:ab,ti OR lactate:ab,ti OR 'lactic acid':ab,ti

309723 records

**Genetics**

#16 'genome-wide association study'/exp OR 'single nucleotide polymorphism'/exp OR 'genetic variation'/exp OR 'genetic risk score'/exp OR 'microrna'/exp OR 'genetics'/exp OR 'genomics'/exp OR genetics*:ab,ti OR genom*:ab,ti OR 'genome-wide association study':ab,ti OR gwas:ab,ti OR 'single nucleotide variation':ab,ti OR 'single nucleotide polymorphism':ab,ti OR snp:ab,ti OR 'genetic variation':ab,ti OR 'genetic risk score':ab,ti OR 'genotype risk score':ab,ti OR grs:ab,ti OR 'polygenic risk score':ab,ti OR 'polygenic score':ab,ti OR prs:ab,ti OR 'micro rna':ab,ti OR mirna:ab,ti

2236966 records

**Epigenetics**

#17 'cpg island'/exp OR 'dna methylation'/exp OR 'methylome'/exp OR 'epigenetics'/exp OR epigenetics:ab,ti OR 'cpg ilands':ab,ti OR 'dna methylation':ab,ti OR epigenomes:ab,ti OR methylome:ab,ti

170397 records

**Exosomes**

#18 'exosome'/exp OR exosome*:ab,ti
43335 records

**All biomarkers combination**
#19 #7 OR #8 OR #9 OR #10 OR #11 OR #12 OR #13 OR #14 OR #15 OR #16 OR #17 OR #18

12968907 records

#20 age:ab,ti OR ethnic*:ab,ti OR white:ab,ti OR caucasian:ab,ti OR asian:ab,ti OR african:ab,ti OR social:ab,ti OR economic:ab,ti OR socioeconomic:ab,ti OR socio-economic:ab,ti

5527104 records

#21 'exercise'/exp OR 'physical activity'/exp

795039 records

#22 exercise:ab,ti OR physical activity:ab,ti

340721 records

#23 #21 OR #22

954221 records

#24 ‘body mass’/exp OR 'body composition'/exp OR 'anthropometry'/exp OR 'obesity'/exp

1058932 records

#25 ‘BMI’:ab,ti OR ‘bodymass’:ab,ti OR ‘body mass’:ab,ti OR ‘body mass index’:ab,ti OR 'body composition':ab,ti OR anthropometry:ab,ti OR obesity:ab,ti OR overweight:ab,ti OR obes*:ab,ti OR 'excess body weight':ab,ti OR 'excess body fat':ab,ti OR 'excess weight':ab,ti OR 'body weight':ab,ti OR lean:ab,ti OR fat:ab,ti OR 'muscle mass':ab,ti

1405134 records

#26 #24 OR #25

1696955 records

#27 'diet therapy'/exp OR 'diet'/exp

704652 records

#28 (diet:ab,ti OR food:ab,ti OR nutriti*:ab,ti OR 'diet therapy':ab,ti OR 'diet intervention':ab,ti OR eating habit*:ab,ti) AND (composition:ab,ti OR pattern*:ab,ti OR matrix*:ab,ti OR 'percent calories':ab,ti)

11381 records

#29 #27 OR #28

711586 records

#30 'health behavior'/exp

459660 records

#31 'behavio* health':ab,ti OR behavi*:ab,ti

1623947 records

#32 #30 OR #31

1967208 records

**Combination phenotype/biomarkers/age-ethnicity-social/lifestyle factors**

#33 #6 OR #19 OR #20 #23 OR #26 OR #29 OR #32

19415016 records

**Subclassification block**

#34 'subtype*':ab,ti OR 'subclassif*':ab,ti OR 'stratif*':ab,ti OR 'subgroup*':ab,ti

1033219 records

**Risk block/prediction block RQ1**

#35 prognos*:ab,ti OR progress*:ab,ti OR predict*:ab,ti OR model*:ab,ti OR nomogram:ab,ti OR statistical:ab,ti OR score*:ab,ti OR risk*:ab,ti OR 'risk score':ab,ti OR 'risk categor*':ab,ti OR 'risk factor*':ab,ti OR 'risk assessment':ab,ti OR algorithm*:ab,ti OR equation*:ab,ti OR precision:ab,ti OR personali*:ab,ti

11779846 records.

#36 #34 OR #35

12001590 records

**Perinatal outcomes block**

#37 'birth weight'/exp OR 'large for gestational age'/exp OR 'small for date infant'/exp OR 'cesarean section'/exp OR 'neonatal respiratory distress syndrome'/exp OR 'neonatal respiratory failure'/exp OR 'hypoglycemia'/exp OR 'polycythemia'/exp OR 'macrosomia'/exp OR 'congenital malformation'/exp OR 'birth injury'/exp OR 'hyperbilirubinemia'/exp OR 'perinatal mortality'/exp OR 'perinatal morbidity'/exp OR 'newborn mortality'/exp OR 'newborn morbidity'/exp OR 'fetus death'/exp

1429308 records

#38 (perinatal:ab,ti OR pregnancy:ab,ti OR neonatal:ab,ti OR fetal:ab,ti OR foetal:ab,ti) AND outcome*:ab,ti OR birthweight:ab,ti OR 'birth weight':ab,ti OR 'large for gestational age':ab,ti OR lga:ab,ti OR 'small for gestational age':ab,ti OR 'small-for-gestational-age':ab,ti OR sga:ab,ti OR preterm:ab,ti OR 'cesarean delivery':ab,ti OR cs:ab,ti OR 'neonatal respiratory morbidity':ab,ti OR 'neonatal respiratory stress':ab,ti OR hypoglycemi*:ab,ti OR polycythemi*:ab,ti OR macrosomi*:ab,ti OR 'congenital anomal*':ab,ti OR 'congenital malformation* birth injur*':ab,ti OR hyperbilirubinemia:ab,ti OR ((intrauterine:ab,ti OR fetal:ab,ti OR foetal:ab,ti OR foetus:ab,ti OR fetus:ab,ti) AND death:ab,ti) OR ((perinatal:ab,ti OR neonatal:ab,ti) AND (mortality:ab,ti OR morbidity:ab,ti)) OR (diagnostic AND zone*:ab,ti) OR 'lipocalin 2':ab,ti

648426 records

#39 #37 OR #38

1797424 records

**Combination search RQ1A** (GDM AND phenotype/biomarkers/age-ethnicity-social/lifestyle factors AND stratification/risk AND perinatal outcomes)

#40 #3 AND #33 AND #36 AND #39

14535 records

AND [embase]/lim NOT ([embase]/lim AND [medline]/lim)

6388 records

Filter Human, English

5600 records

NOT 'conference abstract':it

1705 records

NOT ‘review’:it

1319 records

**Q1**
**PubMED**
**#**1
"Diabetes, Gestational"[Mesh]
=15181

#2
"gestational diabetes"[Title/Abstract] OR GDM[Title/Abstract] OR CGM[Title/Abstract] OR pregnancy induced diabetes[Title/Abstract] OR pregnancy-induced diabetes[Title/Abstract]
=21947

#3
#1 OR #2
=25718

**2022-03-04**
**1A**
**BMI, Biomarkers, age, ethnicity, sociocultural factors, behavioral factors, diet, exercise**

#4
((((((((((((("Phenotype"[Mesh]) OR "Genetic Heterogeneity"[Mesh]) OR "Insulin Resistance"[Mesh]) OR "Genomics"[Mesh]) OR "Metabolomics"[Mesh]) OR "Proteomics"[Mesh]) OR "Lipidomics"[Mesh]) OR "MicroRNAs"[Mesh]) OR "Lipids"[Mesh]) OR "Triglycerides"[Mesh]) OR "Adipokines"[Mesh]) OR "Leptin"[Mesh]) OR "Adiponectin"[Mesh]) OR "Pregnancy-Associated Plasma Protein-A"[Mesh] OR "biomarkers"[Mesh]) OR "exosomes"[Mesh])

=2554133

#5

Sex hormone-binding globulin[Title/Abstract] OR SHBG[Title/Abstract] OR estrogen[Title/Abstract] OR cortisol[Title/Abstract] OR prolactin[Title/Abstract] OR progesterone[Title/Abstract] OR insulin-like growth factor[Title/Abstract] OR IGF[Title/Abstract] OR placenta-derived[Title/Abstract] OR human placental lactogen[Title/Abstract] OR hPL[Title/Abstract] OR human chorionic somatomammotropin[Title/Abstract] OR HCS pregnancy associated plasma protein A[Title/Abstract] OR PAPP-A[Title/Abstract] OR Placental growth hormone[Title/Abstract] OR PGH[Title/Abstract] OR Thyroid*[Title/Abstract] OR TSH[Title/Abstract] OR thyroglobulin antibody[Title/Abstract] OR thyroid peroxidase antibody[Title/Abstract] OR "free T4"[Title/Abstract] OR "free T3"[Title/Abstract] OR Metabolomics*[Title/Abstract] OR butyrate[Title/Abstract] OR butyric acid[Title/Abstract] OR acetylcarnitine[Title/Abstract] OR acylcarnitine[Title/Abstract] OR carnitine[Title/Abstract] OR lactate[Title/Abstract] OR lactic acid[Title/Abstract] OR Genome-Wide Association Study[Title/Abstract] OR GWAS[Title/Abstract] OR single nucleotide variation[Title/Abstract] OR Polymorphism, Single Nucleotide[Title/Abstract] OR SNP[Title/Abstract] OR genetic variation[Title/Abstract] OR genetic risk score[Title/Abstract] OR genotype risk score[Title/Abstract] OR GRS[Title/Abstract] OR polygenic risk score[Title/Abstract] OR polygenic score[Title/Abstract] OR PRS[Title/Abstract] OR CpG

Islands[Title/Abstract] OR DNA Methylation[Title/Abstract] OR Epigenomes[Title/Abstract] OR Methylome[Title/Abstract]

=950352

#6

lipids OR triacylglycerol*[Title/Abstract] OR triglyceride*[Title/Abstract] OR VLDL[Title/Abstract] OR very low density lipoprotein[Title/Abstract] OR cholesterol OR lipoprotein* OR hdl[Title/Abstract] OR ldl[Title/Abstract] OR hyperlipidemia* OR apolipoprotein*[Title/Abstract] OR non-hdl[Title/Abstract] OR lipidemia*[Title/Abstract] OR lipemia*[Title/Abstract] OR Lipemic[Title/Abstract] OR dyslipidemia OR fatty acid[Title/Abstract] OR Amino Acid* OR aminoacid* OR bcaa[Title/Abstract] OR creatine[Title/Abstract] OR leucine[Title/Abstract] OR isoleucine[Title/Abstract] OR valine[Title/Abstract] OR alanine[Title/Abstract] OR glutamine[Title/Abstract] OR glutamate[Title/Abstract] OR betaine[Title/Abstract] OR creatine [Title/Abstract] OR aspartate[Title/Abstract] OR tyrosine[Title/Abstract] OR glutathione[Title/Abstract] OR serine[Title/Abstract] OR threonine[Title/Abstract] OR histidine[Title/Abstract] OR tryptophan[Title/Abstract] OR glycine[Title/Abstract] OR inflammation OR Cytokine*[Title/Abstract] OR interluekin*[Title/Abstract] OR IL[Title/Abstract] OR CRP[Title/Abstract] OR high-sensitivity CRP[Title/Abstract] OR hs-CRP[Title/Abstract] OR TNF-alpha OR Tumor Necrosis Factor alpha OR MCP-1 OR Monocyte Chemoattractant Protein-1 OR chemokine OR adipokines[Title/Abstract] OR adipocytokines[Title/Abstract] OR adiponectin[Title/Abstract] OR leptin[Title/Abstract] OR RBP4[Title/Abstract] OR retinol binding protein 4[Title/Abstract] OR exosome*[Title/Abstract]=4363686

#7
#4 OR #5 OR #6
=5991911

#8
age[Title/Abstract] OR ethnic*[Title/Abstract] OR white[Title/Abstract] OR caucasian[Title/Abstract] OR asian[Title/Abstract] OR african[Title/Abstract] OR american[Title/Abstract]
=3322327

#9
"Exercise"[Mesh] OR exercise[Title/Abstract] OR physical activit*[Title/Abstract]
=484766

#10
BMI[Title/Abstract] OR body mass[Title/Abstract] OR bodymass[Title/Abstract] OR "body mass index"[Title/Abstract] OR body composition[Title/Abstract] OR anthropometr*[Title/Abstract] OR obes*[Title/Abstract] OR overweight[Title/Abstract] OR "over weight"[Title/Abstract] OR excess body weight[Title/Abstract] OR excess body fat[Title/Abstract] OR excess weight[Title/Abstract] OR body weight[Title/Abstract] OR bodyweight[Title/Abstract] OR lean[Title/Abstract] OR fat[Title/Abstract] OR muscle mass[Title/Abstract] OR musclemass[Title/Abstract]

=1001815

#11
(("Diet Therapy"[Mesh]) OR "Diet"[Mesh]) OR ((diet[Title/Abstract] OR food[Title/Abstract] OR nutriti*[Title/Abstract] OR dietary[Title/Abstract] OR "eating habit*"[Title/Abstract]) AND (composition*[Title/Abstract] OR pattern*[Title/Abstract] OR matrix*[Title/Abstract] OR "percent calories"[Title/Abstract]))
=433783

#12
(“Health behavior"[Mesh]) OR ((behavio*[Title/Abstract]) AND (health*[Title/Abstract]))
=589285

#13
#7 OR #8 OR #9 OR #10 OR #11 OR #12
=9842933

**Subclassification**

#14
subtype*[Title/Abstract] OR subclassif*[Title/Abstract] OR stratif*[Title/Abstract] OR subgroup*[Title/Abstract]
=694723

**Risk/Prediction**

#15
prognos*[Title/Abstract] OR progress*[Title/Abstract] OR predict*[Title/Abstract] OR model*[Title/Abstract] OR nomogram*[Title/Abstract] OR statistical[Title/Abstract] OR score*[Title/Abstract] OR risk categor*[Title/Abstract] OR risk factor*[Title/Abstract] OR risk assessment[Title/Abstract] OR algorithm*[Title/Abstract] OR equation*[Title/Abstract] OR precision[Title/Abstract] OR personali*[Title/Abstract]
=7803055

#16
#13 OR #14
=8128024

**Perinatal outcomes**

#17
Perinatal outcome*[Title/Abstract] OR pregnancy outcome*[Title/Abstract] OR "large for gestational age"[Title/Abstract] OR LGA[Title/Abstract] OR birthweight[Title/Abstract] OR small-for-gestational-age SGA preterm*[Title/Abstract] OR "cesarean deliver*"[Title/Abstract] OR large-for-gestational-

age[Title/Abstract] OR neonatal respiratory morbidity[Title/Abstract] OR neonatal respiratory stress[Title/Abstract] OR hypoglycemia[Title/Abstract] OR hypoglycaemia[Title/Abstract] OR polycythemia[Title/Abstract] OR macrosomi*[Title/Abstract] OR "birth injur*"[Title/Abstract] OR hyperbilirubinemia[Title/Abstract] OR congenital malformations[Title/Abstract] OR intrauterine death[Title/Abstract] OR fetal outcome*[Title/Abstract] OR perinatal mortality[Title/Abstract] OR perinatal morbidit*[Title/Abstract]
=159004

#18
((("Hypoglycemia"[Mesh]) OR "Polycythemia"[Mesh]) OR "Respiratory Distress Syndrome, Newborn"[Mesh]) OR "Perinatal Death"[Mesh]
=53188

#19
#17 OR #18
=186624

**Combined**

#20
#3 AND #13 AND #16 AND #19
=3497

#21
#20 Filters: Humans, English
=**2861 references**

**#22**
(review[Title/Abstract]) OR (review[Publication Type])
=3665456

#23
#21 NOT #22
=**2442 references**

#### Supplementary Material 2: Modified Joanna Briggs Institute critical appraisal checklist for cohort studies

**Modified JBI Critical Appraisal Checklist**

This form captures several categories of factors that could induce bias 1) selection, 2) confounding, 3) the outcome was not present at the time of exposure assessment (eg, time of precision variable assessment), 4) loss to follow-up

---

**Selection**

The following questions are used to assess whether the method of selecting GDM cases, GDM subtypes, or GDM precisions variables may have resulted in biased findings or conclusions. If there are potential biases, then consider carefully whether the study should be included.

**1a) Sample size.**

Are there enough GDM cases in each subtype, or with the precision variable measured, for us to draw reliable conclusions. For instance, potentially 30-50 in each GDM group?

*Check all that apply.*

O Yes O No O Unclear O Not applicable

*1b) Optional: supporting text or note*

**2a) Are both subtypes selected from the same community/population; or if the subtype is sociocultural, are the subtypes selected with the same criteria.** (This is unlikely to occur frequently)

*Check all that apply.*

O Yes O No O Unclear O Not applicable

*2b) Optional: supporting text or notes*

**3a) Were the subtypes measured similarly in all women.**

Highest quality is if the same method is used for all participants.

*Check all that apply.*

O Yes O No O Unclear O Not applicable

*3b) Optional: supporting text or notes*

**4a) Were the subgroups ascertained in a reliable way (eg, hospital record, interview).**

Secure record or structured interview are considered highest quality. Written self-report or medical record are considered mediocre quality. No description is poor quality.

*Check all that apply.*

O Yes O No O Unclear O Not applicable

*4b) Optional: supporting text or notes*

**Confounding**

The purpose of these questions is to help guide us to think about whether we can draw valid conclusions from the study, or whether other factors (confounders) could be influencing the findings.

**5a) Were confounding factors identified.**

a confounder is a variable that influences both the exposure variable and outcome variable, that isn't on the causal pathway and results in a spurious association

*Check all that apply.*

O Yes O No O Unclear O Not applicable

*5b) Optional: supporting text or notes*

**6a) Were strategies to deal with confounding stated.**

Adjusted analyses, or subgroup analyses could count. For instance, the study controls for BMI (or other potential confounders) when comparing outcomes between subtypes

*Check all that apply.*

O Yes O No O Unclear O Not applicable

*6b) Optional: supporting text or notes*

**Outcome**

The purpose of this section is to assess whether the individuals had the outcome when the subtype was measured. We are trying to assess/establish whether temporality between exposure and outcome is well defined.

**7a) Were the groups free of the outcome at the time of subtype measurement.**

The goal of this question is to assess whether the subtype or precision variable influences the outcome. Said differently, the goal is to assess temporality of exposure and outcome (even though we understand that sometimes the pathophysiology of the outcomes we are interested in begin in early pregnancy).

*Check all that apply.*

O Yes O No O Unclear O Not applicable

*7b) Optional: supporting text or note*

**8a) Were the outcomes measured in a valid and reliable way.**

If there are multiple outcomes and not all are measured well, please make a note. Overall, the goal is to help you determine whether the study should be included.

*Check all that apply.*

O Yes O No O Unclear O Not applicable

*8b) Optional: supporting text or notes*

**Loss to follow-up**

If there was substantial loss to follow-up (eg, >25%), then are the women lost inherently different and could this result in systematic differences in outcomes. The goal is to help us think about whether the loss to follow-up results in a lower quality study that we would be unable to draw valid conclusions from.

**9a) Was there adequate follow up of all participants.**

The goal is to assess whether any bias could be introduced due to loss to follow-up. High quality is complete follow up (all accounted for), or the % lost to follow up is unlikely to introduce bias, or there was a description of those lost.

*Check all that apply.*

O Yes O No O Unclear O Not applicable

*9b) Optional: supporting text or notes.*

**10a) Was appropriate statistical analysis used.**

*Check all that apply.*

O Yes O No O Unclear O Not applicable

*10b) Optional: supporting text or notes.*

**11a) Overall appraisal.**

Please pick exclude if we should not summarize OR meta analyze the study.

*Check all that apply.*

O Yes O No

**12a) Overall quality.**

*Check all that apply.*

O High O Low

*11a-12a) Optional: supporting text or notes of overall appraisal.*
